# Insights into the relationship between nasal bacterial composition and susceptibility to early-life respiratory disease: a pilot observational study

**DOI:** 10.1101/2025.08.16.25333459

**Authors:** Jose A. Caparrós-Martín, Elizabeth Kicic-Starcevich, Patricia Agudelo-Romero, David G Hancock, Thomas Iosifidis, Yuliya V Karpievitch, David J Martino, Guicheng Zhang, Desiree T Silva, Anthony Bosco, Susan L Prescott, Peter N LeSouëf, Anthony Kicic, Stephen M. Stick

## Abstract

**Background:** Early-life susceptibility to viral respiratory infections is associated with long-term respiratory morbidity in children. Currently, no reliable tools exist to predict susceptibility to these infections. Because the endogenous microbiota can influence pathogen virulence and airway inflammation, it represents a potential target for prevention. In this pilot study, we hypothesised that distinct nasal microbial communities are associated with susceptibility to viral respiratory infections and wheezing outcomes during the first year of life.

**Methods:** We analyzed 90 nasal swabs from 55 infants enrolled in the AERIAL study, representing asymptomatic samples collected at scheduled visits (∼4 months) and symptomatic samples. Bacterial profiling was done blind to clinical data using full-length *16S rRNA* sequencing, and bacterial load was quantified using a pan-bacterial TaqMan® assay.

**Results:** Bacterial diversity was similar between asymptomatic and symptomatic swabs, with community composition differences largely driven by inter-individual variation. In paired samples, symptomatic swabs showed reduced diversity but no change in bacterial load. Virus-bacteria interactions were observed in rhinovirus-positive swabs, but not SARS-CoV-2-positive swabs. Two nasal endotypes were identified, dominated by *Moraxella* or *Streptococcus* and differing in alpha diversity. Endotypes were associated with age at wheeze onset, and their relationship with wheezing episode rates showed a suggestive sex-dependent pattern that warrants further investigation.

**Conclusion:** Our pilot data suggest that the nasal microbiota might shape early wheezing outcomes in a sex-dependent manner, and highlight the value of longitudinal studies for clarifying how host-bacteria-virus interactions in early life.

**What is tested:** Whether distinct nasal microbiota endotypes in infancy are associated with later respiratory phenotypes, particularly wheeze.

**What the study showed:** Two endotypes were identified. Moraxella-dominated and Streptococcus-dominated, with differences in alpha diversity and associations with age at wheeze onset and wheezing rates.

**The significance and clinical impact of the research outcomes:** Nasal microbiota endotypes may help identify infants at risk of wheeze and support early risk stratification, pending validation in larger cohorts.

## Introduction

Acute respiratory infections are a leading cause of death in children under five and a significant source of morbidity, particularly among newborns and infants (1). Most symptomatic viral respiratory infections occur early in life, with recurrent infections associated with an increased risk of developing chronic wheezing disorders during childhood (2, 3). Wheezing disorders including recurrent wheeze and asthma, are among the most prevalent pediatric respiratory illnesses (4). Rates and severity are highest in vulnerable populations, such as Indigenous children, lower-socioeconomic backgrounds, or children with underlying chronic diseases (4). Despite this burden, no clinically validated biomarkers exist to reliably predict which children will develop severe or persistent respiratory disease (4).

Longitudinal studies have shown that nasopharyngeal colonization trajectories influence the risk of respiratory infections during infancy and the later development of asthma and related wheezing disorders (5,6). In early life, the nasopharyngeal microbiota transitions from a community dominated by maternal *Staphylococcus* species to one enriched with *Corynebacterium*, *Dolosigranulum*, or *Moraxella*, with transient colonization by *Haemophilus* or *Streptococcus* (6,7). Early acquisition of stable, pathogen-dominated communities has been linked to airway inflammation and increased respiratory morbidity (5,6,8–12). For example, a recent study found that profiles dominated by *Haemophilus* and *Moraxella* are associated with early interferon-related epithelial responses and pro-inflammatory airway environments (5), whereas *Corynebacterium*-enriched communities correlated with respiratory health (5,6). However, findings across studies vary, with some reporting that *Moraxella*-dominated communities are associated with fewer upper respiratory tract infections and greater protection against viral infections in both adults and children (13–15). These discrepancies may stem, in part, from the limitations of targeting fragments of the *16S rRNA* gene, which do not provide sufficient resolution to determine species-level taxonomy (16).

While the nasopharyngeal microbiota has been well studied, it remains unclear whether nasal microbial communities, which are at the frontline of viral exposure, are associated with viral infection outcomes. We used full-length *16S rRNA* sequencing to profile nasal bacterial communities associated with nasal swabs in the AERIAL birth cohort (17) and elucidate whether specific bacterial communities are associated with susceptibility to symptomatic respiratory infections and respiratory phenotypes in the first year of life. We hypothesised that symptomatic viral respiratory episodes are accompanied by shifts in nasal bacterial community structure, and that distinct baseline nasal community types are associated with subsequent susceptibility to symptomatic infection and wheezing outcomes in the first year of life. As a pilot substudy, we aimed to estimate effect sizes and identify candidate host–microbe–virus patterns for validation in larger longitudinal datasets.

## Methods

Detailed descriptions of the sample, inclusion criteria, DNA extraction protocol, and statistical analyses are provided in the Supplementary Methods.

### AERIAL study cohort

We profiled the nasal microbiota of infants enrolled in the Airway Epithelium Respiratory Illnesses and Allergy (AERIAL) birth cohort study (17), a sub-study nested with the ORIGINS birth cohort (18,19). Respiratory virus data for the first year of life was determined by nasal swabs at scheduled asymptomatic visits (approximately 3, 6, and 9 months of age) and during symptomatic episodes. AERIAL used a smartphone app for real-time monitoring of respiratory symptoms and symptomatic swab collection (17, 20). For this pilot substudy, we selected and analysed the microbiota in a subset of AERIAL nasal swabs (Figure S1), blinded to clinical data. Definitions of symptomatic swabs are provided in supplemental methods. Clinical data (demographics, medical history, illness events, wheeze, allergies and respiratory medications) were collected under AERIAL and ORIGINS protocols (17,18).

### Study size

This was a pilot, exploratory study designed to examine associations and estimate effect sizes for future work. No formal power calculation was performed.

Instead, we analysed a subset of participants randomly selected from the cohort at the time, to ensure feasibility within available resources while maintaining representativeness of the broader population.

### Sample collection

Both nostrils were sampled at scheduled visits and during symptomatic episodes recorded through the smartphone app. One swab was tested with a qPCR respiratory virus panel at a pathology service, and the second swab was preserved for microbiome analyses (17). Additional details are provided in the supplemental methods.

### Microbial DNA extraction, sequencing and data processing

The detailed protocol is shown in the supplemental methods. Briefly, bacterial DNA was extracted from nasal swabs using the QIAamp DNA kit (QIAGEN) incorporating a bead-beating pre-processing step as described previously (21). Negative extraction controls and bacterial load quantification included to identify reagent contaminants (22), as we described (23). The full-length *16S rRNA* gene was amplified (Figure S2), and libraries prepared using the SMRTbell® preparation kit 3.0. Sequencing was carried out on a PacBio Sequel IIe platform. Reads were processed and assigned to taxonomy with DADA2 (v1.28.0) (24), as implemented in the nfcore pipeline ampliseq (v2.8.0), using Nextflow (v25.04.2) (25), the naïve Bayesian classifier implemented in *DADA2* (26), and the Genome Taxonomy Database (vR09-RS220) as reference (27). Guided by negative controls and bacterial load quantification (Figure S3)(28), we identified and removed potential contaminants using the R package *decontam* (v1.20.0) (29). Procrustes analysis confirmed that filtering did not significantly alter the structure of the original taxonomic data (Table S3, Figure S4).

### Statistical analysis

Statistical analyses are comprehensively described in supplemental material. Briefly, analyses were done in R statistical software (v4.3.0) within the RStudio environment (v2023.03.0). Microbiome community structure was evaluated using PERMANOVA on Aitchison distances, with PCA and diversity indices calculated after centered log-ratio transformation using the functions contained in the MixOmics package (v6.24.0) (30). Community clustering was assessed using Dirichlet Multinomial Mixtures (DMM) models (31). Differential abundance was tested with ANCOM-BC2 (32). Additional analyses included correlation tests, linear and logistic regression, and generalized linear models, with false discovery rate correction for multiple testing. Significance was set at *p*<0.05.

### Results Study cohort

In this study, we selected a total of 90 nasal swabs from AERIAL. Figure S1 depicts the sampling structure and clinical metadata is summarised in Table 1 and Tables S4-S5. These swabs were collected from 55 infants (30/54, 54% female) and comprised 42 scheduled (asymptomatic) swabs and 48 symptomatic-episode swabs. Asymptomatic swabs were collected at 3-month (n=25, 59.5%), 6-month (n=15, 35.7%), or 9-month (n=2, 4.8%) visits (median age [IQR]: 131 days [106–199]). The median age for symptomatic swabs was 189 days (IQR [148-247]). Paired samples, consisting of an asymptomatic swab and the first subsequent symptomatic episode, were available for 33 infants. The median interval between paired asymptomatic and symptomatic swabs was 40.5 days (IQR 26.5-62.5).

**Table 1.** Patient demographics and swab characteristics. HMPV, human metapneumovirus; RSV, respiratory syncytial virus. *, four SARS-CoV-2–positive swabs also tested positive for other viruses. Similarly, four rhinovirus-positive swabs and one parainfluenza-positive swab were co-infected with additional viruses. All adenovirus-positive swabs were detected in combination with other viruses. The difference in sample numbers across ages reflects the fact that the study was ongoing at the time of analysis, resulting in more swabs collected at earlier visits.

| Characteristics | Total number of infants n=55 |
| --- | --- |
| ***Sex, n (%)*** |  |
| Male | 25 (46%) |
| Female | 30 (54%) |
| ***Nasal swabs type, n (%)*** |  |
| Scheduled visits (asymptomatic) | 42 (46%) |
| Symptomatic swabs | 48 (54%) |
| ***Asymptomatic swabs age (months), n (%)*** |  |
| 3 months of age | 25 (59.5%) |
| 6 months of age | 15 (35.7%) |
| 9 months of age | 2 (4.8%) |
| ***Viral detection by swab type, n (%)*** |  |
| Scheduled visits (asymptomatic) | 2 (4.8%) |
| Symptomatic swabs | 48 (98%) |
| ***Number of swabs by virus detected, n (%)*** |  |
| Rhinovirus * | 28 (56%) |
| SARS-CoV-2 * | 23 (46%) |
| Adenovirus * | 2 (4%) |
| *Parainfluenza \** | 2 (4%) |
| *HMPV* | 1 (2%) |
| *RSV* | 1 (2%) |

In our cohort, 50 swabs tested positive for respiratory viruses by qPCR, comprising 2/42 asymptomatic swabs and 48/48 symptomatic swabs (Table 1, Figure S1). Of the virus-positive asymptomatic swabs, one was positive for rhinovirus and one for SARS-CoV-2 (Table 1 and Table S4). Among symptomatic swabs, the most frequently detected viruses were rhinovirus (28/48, 56%) and SARS-CoV-2 (23/48, 46%) (Table 1 and Table S5). Five symptomatic swabs from five infants showed polyviral infections: SARS-CoV-2 and rhinovirus in three swabs; SARS-CoV-2, rhinovirus, and adenovirus in one swab; and rhinovirus, adenovirus, and parainfluenza in one swab (Table S5).

### Characterization of the swab-associated nasal bacterial communities

We obtained reliable microbial profiles from 71 (∼79%) swab samples, of which 37 (∼52%) represented asymptomatic swabs. Consistent with previous Western Australian cohorts (6,12), nasal swabs contained commensal and opportunistic taxa typical of the infant nasopharynx, including *Moraxella* and *Streptococcus* species (Figure S5). The dominant amplicon sequence variants (ASVs) were assigned to *Moraxella nonliquefaciens* (mean [SD] 22.55% [36.42]; prevalence 36.62%), *Moraxella catarrhalis* (13.71% [28.16]; 43.66%), and *Streptococcus* (12.14% [16.94]; 85.91%).

Some infants harboured taxa consistent with respiratory pathogens (Figure S5). Given the established associations between early colonization by *Moraxella catarrhalis* or *Streptococcus pneumoniae* and increased susceptibility to respiratory infections and adverse respiratory phenotypes (5,9,11,12), we compared their relative abundance between asymptomatic and symptomatic swabs. *Streptococcus pneumoniae* was higher in symptomatic swabs (Wilcoxon rank sum test (WRST) with false discovery rate (FDR) correction (*r*=0.28, *p*=0.03)) (Figure S6), whereas *Moraxella catarrhalis* did not differ (WRST FDR-corrected (*r*=0.14, *p*=0.23)) (Figure S6).

### Interaction between respiratory viruses and bacterial taxa in the nose

Prior studies report virus-associated shift in the nasal microbiota and suggest viral modulation of bacterial pathogen virulence (6,9,33). To explore multilevel interactions, we correlated the three more abundant ASVs, which together accounted for 60% of the cumulative abundance (Figure 1A), with the two most prevalent viruses; rhinovirus and SARS-CoV-2 (Table 1). Rhinovirus detection correlated positively with *Moraxella catarrhalis* (ρ = 0.37, FDR-adjusted *p* = 0.004), and negatively with *Streptococcus* (ρ = −0.47, FDR-adjusted *p* = 0.0002), while SARS-CoV-2 showed no significant correlations with any of the bacterial features analyzed (Figure 1B). Alpha diversity in rhinovirus-positive symptomatic swabs was lower than in symptomatic swabs positive for SARS-CoV-2 (WRST, *p* = 0.021; Figure 1C). Likewise, differential abundance analysis identified *Moraxella catarrhalis* as the only ASV significantly enriched in rhinovirus-infected samples, with a fold change of 7.44 (95% CI: 2.39–23.18; adjusted *p* = 0.01). Conversely, bacterial load in symptomatic swabs did not vary with the virus detected (Figure 1D).

**Figure 1.**
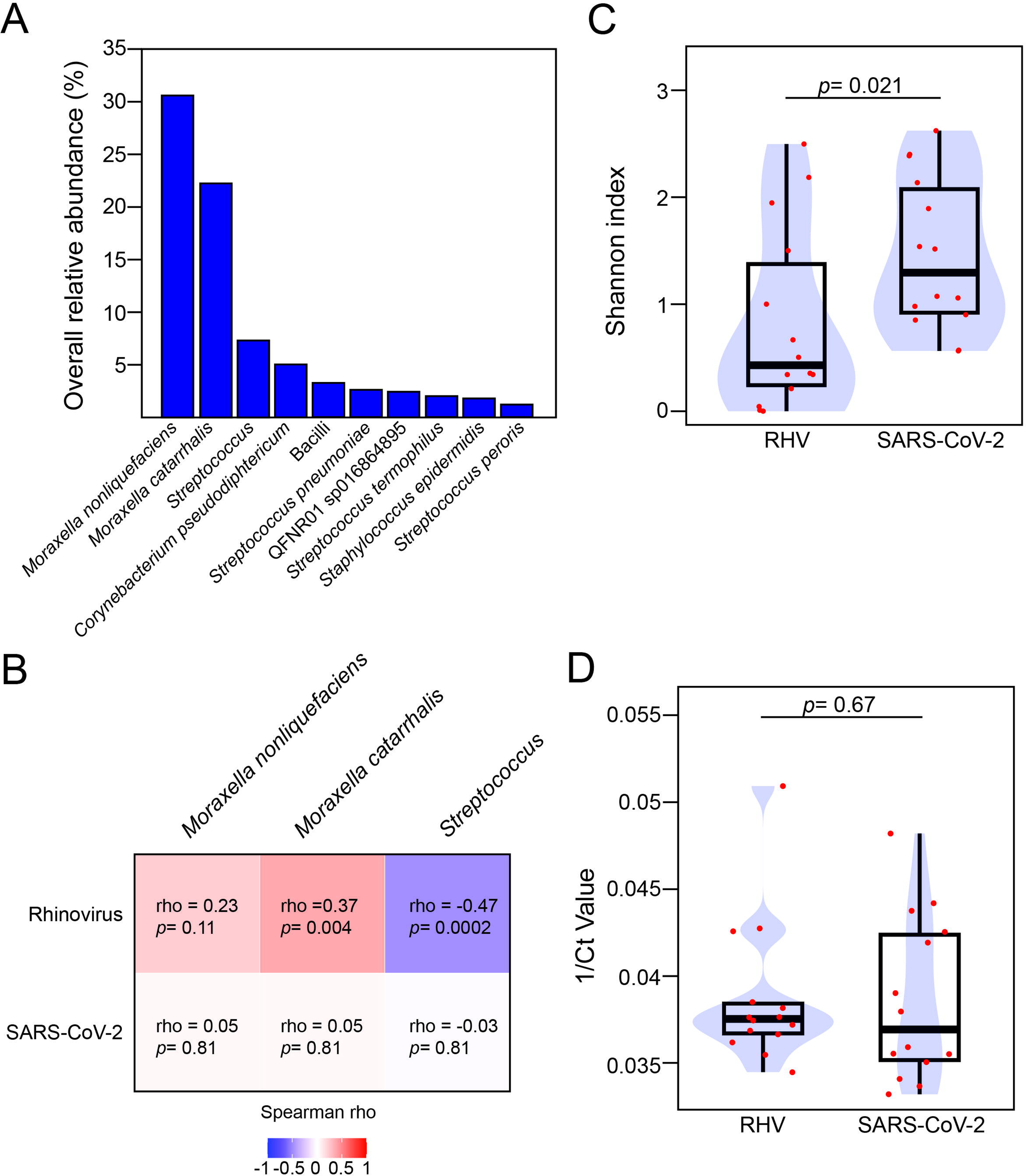
**A.** The bar plot shows the overall relative abundance (%) of the top 10 ASVs in our dataset. **B**. Heatmap summarizing the Spearman correlation analysis between the centered log-ratio transformed counts of the top three ASVs and the two most frequently detected viruses. Heatmap colors indicate the direction of the correlation—positive (red) or negative (blue)—with color intensity reflecting the Spearman’s ρ (rho) coefficient. Both the correlation coefficient and the FDR-adjusted *p*-value are shown within each cell. P-values were corrected for multiple comparisons using the false discovery rate (FDR) method. **C-D**. Boxplots representing alpha diversity (**C**, Shannon index) or bacterial load (**D**) in symptomatic swabs based on the detected virus. RHV: rhinovirus. Boxplots are overlaid with density plots (blue), and individual samples are shown as red dots. P-values for the indicated comparisons were calculated using the Wilcoxon rank-sum test.

### Microbial community analysis across symptomatic and asymptomatic swabs

Next, we evaluated community structural variation between samples collected during scheduled visits and those taken during a symptomatic episode. PERMANOVA with permutations restricted within subject, indicated that swab type (asymptomatic versus symptomatic) explained only a very small fraction of the variation in bacterial community composition (pseudo-*F*=1.13, R²=0.016, *p*=0.002). This small effect size is consistent with the PCA, which showed substantial overlap between groups (Figure S7). Similarly, we did not observe differences in bacterial load or alpha diversity (Figure 2A-B). In contrast, PERMANOVA showed that interindividual differences explained most variance (83%; pseudo-*F*=2.53, R²=0.83, *p*=0.0001), while sex accounted for little variation (pseudo-*F*=1.12, R²=0.016, *p*=1), suggesting that environmental covariates may play a larger role in shaping nasal community composition.

**Figure 2.**
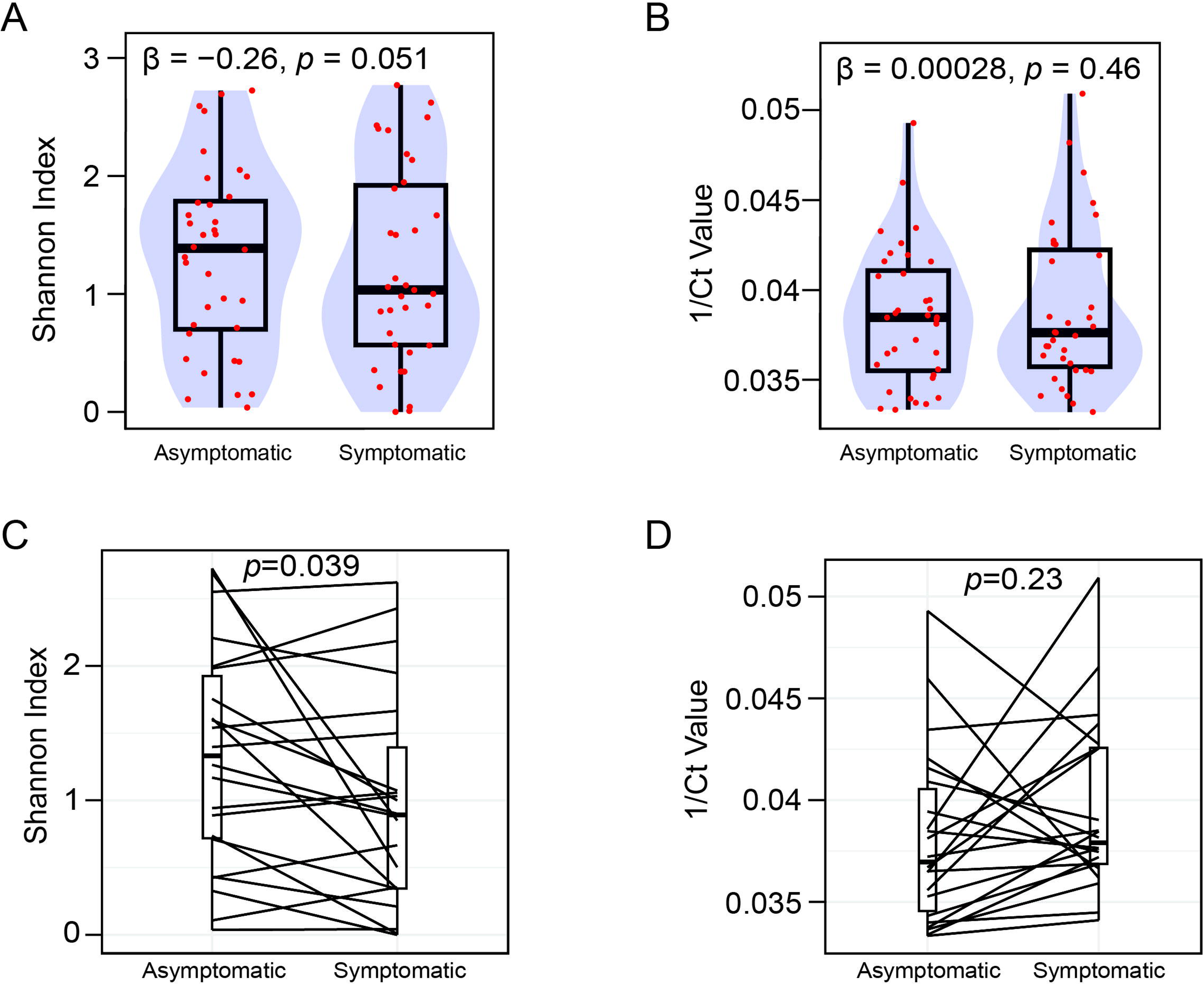
Boxplots representing the bacterial load (**A**, **C**) and alpha diversity ecological parameters (**B**, **D**) associated with DNA extracts from nasal swabs in this study. In panels **C** and **D**, boxplots represent paired nasal swab specimens, with samples obtained from the same infant at scheduled visits (asymptomatic) and during the subsequent symptomatic episode connected by straight lines. For panels **A** and **B**, differences between swab types were tested using linear mixed-effects models with infant as a random intercept to account for repeated measures. Model estimates indicated no difference in bacterial load (β = 0.00028, *p* = 0.46) and no difference in Shannon diversity (β = −0.26, *p* = 0.051).

To account for inter-individual variation, we performed within-infant longitudinal comparisons of asymptomatic and subsequent symptomatic swabs. We observed a reduction in alpha diversity in symptomatic swabs (paired WRST *p*=0.039)), while bacterial load remained unaffected (paired t-test *p*=0.23) (Figure 2C-D).

### Associations between bacterial community structure and future respiratory outcomes

To evaluate whether distinct bacterial community structures were associated with later respiratory outcomes, we modelled the bacterial community data using Dirichlet Multinomial Mixtures (DMM) (31). We identified two microbial community types, hereafter referred to as endotype 1 and endotype 2. Endotype 1 was defined by four ASVs assigned to *Moraxella nonliquefaciens*, *Moraxella catarrhalis*, *Streptococcus*, and *Corynebacterium pseudodiphtheriticum* (Figure 3B and Figure S8). In contrast, endotype 2 was primarily defined by an ASV assigned to the *Streptococcus* genus (Figure 3B and Figure S8). Alpha diversity in endotype 2 was higher than in endotype 1 (WRST (*r*=0.77, *p*=8.56×10^−14^)) (Figure 3C). Neither endotype was associated with sex or seasonality (Figure S9A-B), and endotype assignment in asymptomatic swabs was not associated with childcare attendance (Figure S9C). In asymptomatic swabs, age at sampling was also not associated with endotype assignment (logistic regression, OR per 30 days= 0.87; 95%CI 0.61-1.22; *p*=0.42), including after adjustment for sex (*p*=0.49), or with alpha diversity (Spearman ρ=−0.14, *p*=0.41). Consistent with these findings, age at sampling was not associated with overall community composition in asymptomatic swabs (PERMANOVA, pseudo-*F*=1.58, R²=0.04, *p*=0.14))

**Figure 3.**
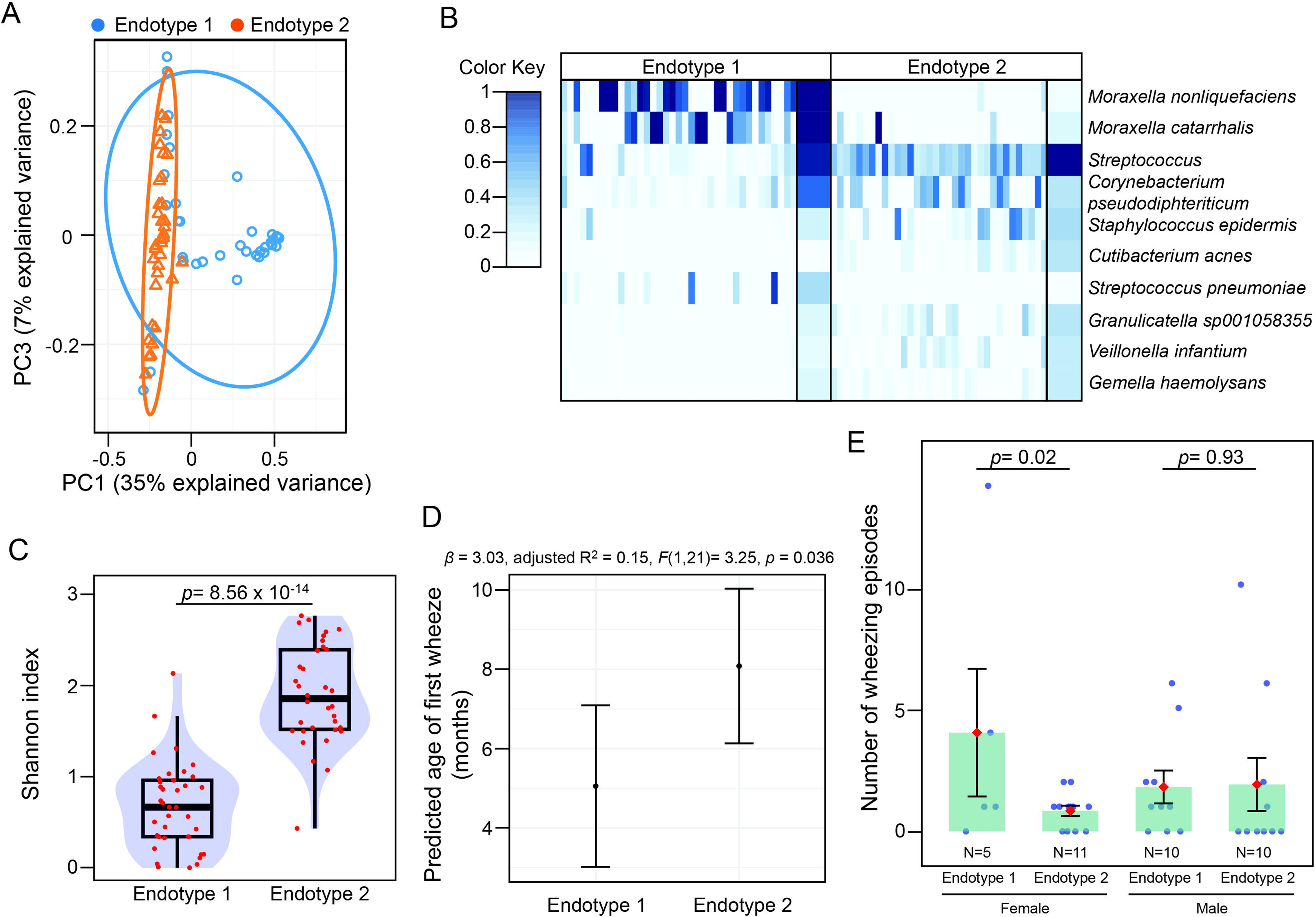
**A.** Principal component analysis illustrating the linear projection of the compositional profiles of the microbiota onto the first and third component of the model, with each sample labelled according to its DMM-model based endotype membership. **B.** Heatmap summarizing the microbial profiles grouped by Dirichlet component. Rows represent the indicated ASVs ordered by their contribution to each component from top to bottom. For simplicity only the 10 top ASVs are represented. The color intensity in the heatmap corresponds to the square root of each ASV’s relative abundance. Narrow columns represent individual swab samples, while wide columns show the mean abundance of each ASV within the Dirichlet components. **C.** Boxplots representing alpha diversity (Shannon index) in relation to the two microbial community types or endotypes represented in (**A**). **D.** Predicted age at first wheeze (in months) with 95% confidence intervals by nasal microbial endotypes (defined from asymptomatic swabs). The estimate endotype effect, adjusted R², and *F*-test results of the fitted linear model are shown at the top of the panel. **E.** Wheezing episode counts shown alongside predictions from the negative binomial model, with endotype (from asymptomatic swabs) as the primary predictor and results stratified by sex. Grey points represent individual observations, while the bars display the mean observed values per endotype-by-sex group, with error bars indicating the standard error of the mean. Red diamonds indicate model-predicted wheezing episodes. Group sample sizes (N) are shown beneath each bar. Estimated marginal means were compared using Wald z-tests with Dunnett adjustment relative to the reference endotype (endotype 1). A sensitivity analysis associated with this panel is shown in supplemental Figure S12.

We evaluated whether nasal microbial endotypes were associated with wheezing outcomes during the first year of life using negative binomial regression. Infants in endotype 1 (reference group) had a mean of 2.53 wheezing episodes (95% CI: 1.33–5.28, *p*=0.007). Compared with endotype 1, infants in endotype 2 had a lower wheezing rate (IRR = 0.52), although this difference was not significant (95% CI: 0.20–1.29, *p*=0.16)(Figure S10A). In addition, infants in endotype 2 had a later onset of wheeze by an estimated 3.03 months compared with endotype 1 (β=3.03, 95% CI: 0.21-5.85, *F*(1, 21)=4.99, *p*=0.036) (Figure 3D).

Building on prior research identifying sex-specific early-life risk factors for wheeze (34), we next investigated whether the association between endotype and wheezing differed by sex. In a negative binomial model including an endotype-by-sex interaction, females in endotype 1 (*Moraxella*-dominated, reference group) had an estimated mean of 4 wheezing episodes (95% CI: 1.52–13.60, *p*=0.011; Figure 3E). Among females, endotype 2 (*Streptococcus*-dominated) was associated with a significantly lower rate compared with endotype 1 (IRR=0.21, 95% CI: 0.05–0.79, *p*=0.026), corresponding to an approximate 79% reduction (Figure 3E, Figure S10B). Within endotype 1, male sex was not significantly associated with wheezing frequency (IRR=0.45, 95% CI: 0.11–1.67, *p*=0.25). The endotype-by-sex interaction suggested that the endotype association may differ by sex, but this pattern was not significant (endotype-by-sex interaction IRR=5.07, 95% CI: 0.86–32.83, *p*=0.079). Consistent with this, the estimated endotype difference among males was close to null (IRR ∼ 1.06). Classification of samples into endotypes using an alternative probabilistic mixed model (Latent Dirichlet Allocation) yielded comparable results to the DMM approach (Figure S11). Overall, the endotype association was driven by females (endotype 2 was associated with lower wheeze rates compared to endotype 1), whereas no clear endotype-associated difference was observed in males.

Notably, we identified a single influential data point with high leverage: a female participant with endotype 1 who experienced 14 wheezing episodes in the first year (Table S4, infant_11). Although this data point is valid and reflects natural variation in our cohort, its removal rendered the association non-significant, indicating sensitivity to individual cases (Figure S12). Thus, our findings suggesting a sex-specific link between the nasal microbiota endotypes and wheezing outcomes should be interpreted with caution and warrant validation in larger, independent datasets.

We found no association between asymptomatic swab endotypes and the number of symptomatic swabs or bronchiolitis episodes during the first year of life (Figure S13A-B), including after adjusting for sex (Figure S13C-D). Similarly, compared to endotype 1, endotype 2 was not associated with the odds of wheezing, recurrent wheezing, or bronchiolitis during the first year of life, with or without adjustment for sex (Figure S14).

To account for interindividual responses, we examined temporal transitions in nasal microbial endotypes between paired asymptomatic and symptomatic swab samples collected and linked these transition patterns to respiratory phenotypes (Figure S15). We observed three types of transitions: from endotype 1 to endotype 1, from endotype 2 to endotype 1, and from endotype 2 to endotype 2. Notably, no participants transitioned from endotype 1 to endotype 2, suggesting stability of endotype 1 during symptomatic episodes. Despite these patterns, we observed no significant differences in the wheezing episodes or symptomatic swab counts across transition groups (Figure S15).

## Discussion

In this observational study, we characterized the nasal microbiota of infants naturally exposed to respiratory viruses during their first year of life in a well-phenotyped infant cohort. We compared the microbiota of asymptomatic and symptomatic swabs to identify microbial patterns associated with respiratory illness and wheeze. We found that nasal bacterial communities were highly variable between infants and more complex than previously described in early-life nasopharyngeal studies. Although overall microbial composition did not differ between asymptomatic and symptomatic swabs, paired longitudinal analyses showed a reduced alpha diversity during symptomatic episodes.

These effects also appeared to differ by virus type, with clearer microbiota associations observed for rhinovirus infections than for SARS-CoV-2. Finally, probabilistic modelling identified two compositional endotypes, with the *Moraxella*-dominated endotype associated with earlier and more frequent wheeze.

The nasal microbiota in our cohort comprised a mixture of commensal and opportunistic taxa, particularly *Streptococcus* and *Moraxella* species, and showed substantial interindividual heterogeneity. This contrasts with previous studies of the infant nasopharynx, which often report simpler communities dominated by a single taxon (5,6,11–13). This difference may reflect both technical and biological factors, including the higher taxonomic resolution of full-length *16S rRNA* gene sequencing and the fact that the anterior nares are likely more exposed to environmental inputs than the nasopharynx. *Streptococcus pneumoniae* was enriched in symptomatic swabs, whereas *Moraxella catarrhalis* was highly prevalent across our cohort but did not differ significantly between asymptomatic and symptomatic swabs. The high prevalence of *Moraxella* species also contrasts with previous research conducted in Perth in which early-life nasopharyngeal colonization by *Moraxella* was not observed (12). This discrepancy may reflect differences in anatomical sampling site, sequencing approach, or cohort characteristics, although broader temporal changes in airway colonization patterns cannot be excluded.

Despite the high viral burden in symptomatic swabs, community-level analyses did not show differences in overall microbial composition, alpha diversity, or bacterial load between asymptomatic and symptomatic samples. This contrasts with previous reports of reduced microbial diversity and increased dominance of potentially pathogenic taxa during viral respiratory infections (6,35,36). We suggest this discrepancy might be due to the high interindividual variability in microbial composition within our cohort and the limited sample size. In paired samples from the same child, however, symptomatic swabs were associated with lower bacterial alpha diversity, suggesting that infection-related alterations in the microbial communities may be more apparent within individuals than across the cohort.

Virus-specific analyses further suggested that not all respiratory viruses interact similarly with nasal bacterial communities. Rhinovirus-positive symptomatic swabs showed lower bacterial alpha diversity than SARS-CoV-2-positive swabs, and rhinovirus detection was associated with enrichment of *Moraxella catarrhalis* and reduced abundance of *Streptococcus*. Differential abundance analysis further revealed a significant enrichment in *Moraxella catarrhalis* in rhinovirus-positive swabs. These findings are consistent with previous reports of rhinovirus- *Moraxella catarrhalis* interactions and support the idea that rhinovirus may preferentially occur in, or promote, a *Moraxella catarrhalis*-enriched airway environment (9,33,37). In contrast, we did not identify clear associations between bacterial taxa and SARS-CoV-2 detection. One possible explanation is that rhinovirus and SARS-CoV-2 differ in the way they interact with the airway environment, potentially reflecting differences in host adaptation and ecological integration, although this remains speculative.

Probabilistic modelling identified two endotypes: a low-diversity, *Moraxella*-dominated endotype (endotype 1) and a *Streptococcus*-dominated endotype (endotype 2). These endotypes were not associated with demographic or environmental variables, but they differed in relation to respiratory outcomes. Infants assigned to the *Streptococcus*-dominated endotype had later wheeze onset than those in the *Moraxella*-dominated endotype. Additionally, the Streptococcus-dominated endotype was associated with fewer wheezing episodes, with this contrast being more evident among females. Although the sex-specific result should be interpreted cautiously, the overall direction is consistent with previous studies linking early *Moraxella*-dominated airway communities with recurrent wheeze and asthma risk (11,38). Notably the *Moraxella*-dominated endotype was characterised not only by *Moraxella catarrhalis*, but also by *Moraxella nonliquefaciens*, suggesting that wheeze-associated *Moraxella* profiles may extend beyond a single species.

Temporal analysis of endotype stability in longitudinal samples revealed that most individuals retained their initial endotype during symptomatic episodes, particularly those with asymptomatic swab communities classified as *Moraxella*-dominated. Transitions were observed from the *Streptococcus*-dominated endotype to the *Moraxella*-dominated endotype, but not the reverse, suggesting that the latter may represent a relatively stable community during illness. Similar stability of *Moraxella*-dominated airway communities has been reported in the nasopharynx (6).

Our pilot study differs from previous research in the field in several key aspects, including full length 16S rRNA sequencing for improved species-level resolution, careful filtering of low biomass samples using bacterial load, and prospective symptom monitoring through a purpose-designed smartphone app. This study also has important limitations. First, this was a single-center pilot study, which may limit generalizability. Second, sampling occurred during a period of altered viral circulation following COVID-19 public health measures in Western Australia. Third, the sample size was modest, and the longitudinal analysis was limited to two consecutive swabs per infant, reducing power to detect more complex associations and increasing sensitivity to outliers. Fourth, microbiome sequencing was performed on a subset of available swabs, limiting time-resolved analysis across the full illness history. Finally, *16S rRNA* amplicon sequencing does not provide functional insight into microbiome-virus interactions.

## Conclusion

In conclusion, this pilot observational study suggests that early-life nasal bacterial endotypes may be associated with wheeze risk, potentially in a sex-specific manner. Although preliminary, the findings support further investigation of nasal microbial profiling as a marker of respiratory disease susceptibility in larger, independent cohorts.

## Ethics approval

The AERIAL study was approved by Ramsey Health Care HREC WA-SA, reference number 1908. Informed consent was obtained from the parents or caregivers of all children participating in the AERIAL study.

## Data availability

The R code is available in the accompanying GitHub repository: https://github.com/jacapmar/MicrobiomeAERIAL. De-identified taxonomy count tables for participants who provided consent for data sharing are available in the Mendeley Data repository (39). Raw sequencing files cannot be made publicly available due to ethical and privacy restrictions related to participant confidentiality and the conditions of the study consent.

## Supporting information

STROBE checklist

## Data Availability

The R code is available in the accompanying GitHub repository: https://github.com/jacapmar/MicrobiomeAERIAL. De-identified taxonomy count tables for participants who provided consent for data sharing are available in the Mendeley Data repository (Caparros-Martin, JA, Kicic-Starcevich E, Agudelo-Romero P, Hancock D, Iosifidis T, Karpievitch Y et al. AERIAL Nasal Microbiome Taxonomy Table. Mendeley Data, V1. doi: 10.17632/vtr3m7tvzc.1). Raw sequencing files cannot be made publicly available due to ethical and privacy restrictions related to participant confidentiality and the conditions of the study consent.

https://data.mendeley.com/datasets/vtr3m7tvzc/1

## Acknowledgments.

We are grateful to all participants in the AERIAL study and their families. Their willingness to adapt to COVID pandemic protocols and self-collect nasal swab samples enabled our study to continue. We acknowledge the AERIAL clinical staff: Bailee Renouf, Amy Greenly, Angela Fuery, Jodie Leslie, Tanya Betty, Suzie Cooper, AERIAL laboratory staff: Ashleigh Heng-Chin, Luke Berry, Courtney Kidd, Minda Amin, and ORIGINS staff at Western Diagnostics Pathology: Fiona Rynne and Allan Albornoz for their continuous support and contribution towards the AERIAL study.

AERIAL is a sub-project of ORIGINS. This unique long-term study, a collaboration between The Kids Research Institute Australia and Joondalup Health Campus, is one of the most comprehensive studies of pregnant women and their families in Australia to date, recruiting 10,000 families over a decade from the Joondalup and Wanneroo communities of Western Australia. We are grateful to all the ORIGINS families who support the project. We would also like to acknowledge and thank the following teams and individuals who have made ORIGINS possible: ORIGINS team; Joondalup Health Campus (JHC); members of the ORIGINS Community Reference and Participant Reference Groups; Research Interest Groups and the ORIGINS Scientific Committee; The Kids Research Institute Australia; City of Wanneroo; City of Joondalup; and Professor Fiona Stanley. ORIGINS has received core funding support from the Telethon Perth Children’s Hospital Research Fund, Joondalup Health Campus, the Paul Ramsay Foundation, and the Commonwealth Government of Australia through the Channel 7 Telethon Trust. Substantial in-kind support has been provided by The Kids Research Institute Australia and Joondalup Health Campus.

S.M.S. received funding from the National Health and Medical Research Council (project grant number NHMRC115648) to support the AERIAL study and is also recipient of a NHMRC Investigator grant (NHMRC 2007725). A.K. is a Rothwell Family Fellow. T.I. is supported by a Stan Perron Charitable Foundation People Fellowship. S.P.A.-R. received funding from the Branchi family. A.B. is supported by the NIH (R21 AI176305-01A1, R01AI099108-11A1).

## Author Contributions

J.A.C.-M., E.K.-S., P.A.-R., D.G.H., T.I., Y.V.K., D.J.M., G.Z., L.T., D.T.S., A.B., S.L.P., P.N.L., A.K. and S.M.S participated in the conceptualization of the study. J.A.C.-M. optimized the extraction protocol and performed the extraction and quantification of the microbial DNA. J.A.C.-M. and P.A.-R. carried out the computational analyses. J.A.C.-M. analyzed the data. E.K.-S. coordinated the logistics of AERIAL study including collection of the swab samples and associated metadata. E.K.-S. coordinated accession to the stored samples and extracted the associated metadata. D.G.H. curated the metadata associated with this specific study. S.M.S. conceived the AERIAL study and obtained the funding to support this study. J.A.C.-M. prepared the figures and the original draft for publication. All the authors participated in critical discussions about the results presented in this manuscript.

