## Supplementary material for "Insights into the relationship between nasal bacterial composition and susceptibility to early-life respiratory disease: a pilot observational study": STROBE checklist

STROBE Statement—Checklist of items that should be included in reports of ***cohort studies***

|  | Item No | Recommendation | Page No |
| --- | --- | --- | --- |
| **Title and abstract** | 1 | (*a*) Indicate the study’s design with a commonly used term in the title or the abstract.  **Addressed. The title identifies the work as a “pilot observational study”, and the abstract also refers to it as a pilot study.** | 1 |
|  |  | (*b*) Provide in the abstract an informative and balanced summary of what was done and what was found  **Addressed. The abstract summarises the background, methods, sample size, main findings, and cautious conclusions.** | 3 |
| Introduction | | | |
| Background/rationale | 2 | Explain the scientific background and rationale for the investigation being reported  **Addressed in the Introduction, which reviews early-life respiratory infections, airway microbiota, and the rationale for focusing on the nasal microbiota.** | 5-6 |
| Objectives | 3 | State specific objectives, including any prespecified hypotheses  **Addressed in the final paragraph of the Introduction. The manuscript states the hypothesis that symptomatic viral respiratory episodes are accompanied by shifts in nasal bacterial community structure, and that distinct baseline nasal community types are associated with later symptomatic infection and wheezing outcomes.** | 5-6 |
| Methods | | | |
| Study design | 4 | Present key elements of study design early in the paper  **Addressed in the Methods, where the study is described as a pilot, exploratory observational study within the AERIAL birth cohort.** | 6,10 |
| Setting | 5 | Describe the setting, locations, and relevant dates, including periods of recruitment, exposure, follow-up, and data collection  **Addressed in the main manuscript and further detailed in the supplement. The study was conducted in the AERIAL birth cohort, nested within ORIGINS, in Western Australia. Samples were collected at scheduled asymptomatic visits (~3, 6, and 9 months) and during symptomatic episodes recorded via smartphone app. Seasonal timing of sampling is provided in Supplementary Tables S4–S5.** | 6,8,16 |
| Participants | 6 | (*a*) Give the eligibility criteria, and the sources and methods of selection of participants. Describe methods of follow-up  **Addressed. Infants were enrolled through the AERIAL cohort; this pilot analysed a randomly selected subset of available participants for feasibility. Follow-up included scheduled visits and app-triggered symptomatic sampling. Definitions of symptomatic swabs are provided in the supplement.** | 6 |
|  |  | (*b*) For matched studies, give matching criteria and number of exposed and unexposed  ***Not applicable. This was not a matched study*** |  |
| Variables | 7 | Clearly define all outcomes, exposures, predictors, potential confounders, and effect modifiers. Give diagnostic criteria, if applicable  **Addressed, although definitions are distributed across the manuscript and supplement. Outcomes included wheezing-related phenotypes, bronchiolitis, and symptomatic swab counts; exposures/predictors included nasal microbiota composition, endotype assignment, and viral detection; sex was assessed as an effect modifier; seasonality, childcare attendance, and age at sampling were also evaluated. Symptomatic swab criteria are explicitly defined in the supplement.** | 6-8 |
| Data sources/ measurement | 8* | For each variable of interest, give sources of data and details of methods of assessment (measurement). Describe comparability of assessment methods if there is more than one group  **Addressed. Sample collection, virus testing, microbiome sample handling, DNA extraction, sequencing, bacterial load quantification, and bioinformatic processing are described in the manuscript and in detail in the supplement.** | 7 |
| Bias | 9 | Describe any efforts to address potential sources of bias  **Addressed. Bias-reduction measures included blinded microbiome profiling, negative extraction controls, bacterial load filtering for low biomass samples, contaminant removal with decontam, and smartphone-based real-time symptom monitoring to reduce recall bias.** | 6-7 |
| Study size | 10 | Explain how the study size was arrived at  **Addressed. The study is explicitly described as a pilot exploratory study with no formal power calculation; sample size was determined by feasibility and resource availability.** | 6 |
| Quantitative variables | 11 | Explain how quantitative variables were handled in the analyses. If applicable, describe which groupings were chosen and why  **Addressed. Quantitative microbiome data were handled as compositional data using CLR transformation and Aitchison distances; count outcomes were analysed using Poisson or negative binomial models depending on dispersion; age at sampling and other continuous variables were analysed using regression or correlation methods as appropriate.** | 7 |
| Statistical methods | 12 | (*a*) Describe all statistical methods, including those used to control for confounding  **Addressed in the manuscript and expanded in the supplement. Analyses included PERMANOVA, PCA, DMM modelling, ANCOM-BC2, regression models, mixed-effects models, and FDR correction. Sex was included as an adjustment variable where indicated.** | 7-12 |
|  |  | (*b*) Describe any methods used to examine subgroups and interactions  **Addressed. Sex-stratified analyses and sex-by-endotype interaction terms were used for wheezing outcomes.** |  |
|  |  | (*c*) Explain how missing data were addressed  **Missing/unanalysable microbiome data are reflected in Supplementary Tables S4–S5 through the variable “Nasal microbiota: Yes/No”, indicating whether a swab yielded a reliable microbial profile. The supplement further states that, after filtering, one sample with only 136 reads was removed from further analyses.** |  |
|  |  | (*d*) If applicable, explain how loss to follow-up was addressed  **Largely not applicable. This was an analysis of available swabs within a cohort framework rather than a conventional follow-up study focused on attrition. Sampling availability and pairing are described in the Results and Supplementary Tables S4–S5.** |  |
|  |  | (*e*) Describe any sensitivity analyses  **Addressed. Sensitivity analyses included outlier removal for wheezing analyses, alternative endotype modelling using latent Dirichlet allocation, and assessment of age at sampling as a potential confounder.** |  |
| Results | | |  |
| Participants | 13* | (a) Report numbers of individuals at each stage of study—eg numbers potentially eligible, examined for eligibility, confirmed eligible, included in the study, completing follow-up, and analysed  **Addressed. The manuscript reports 90 selected nasal swabs from 55 infants, numbers of asymptomatic and symptomatic swabs, and the number of reliable microbial profiles obtained after filtering. Supplementary Tables S4–S5 further specify inclusion/exclusion at the sample level.** | 8 |
|  |  | (b) Give reasons for non-participation at each stage  **The manuscript does not provide a participant non-participation flow in the conventional STROBE sense, but the supplement indicates whether individual swabs did or did not yield reliable microbiome profiles, and the processing workflow explains exclusion due to low bacterial biomass/contamination filtering and one low-read sample.** |  |
|  |  | (c) Consider use of a flow diagram  **None provided** |  |
| Descriptive data | 14* | (a) Give characteristics of study participants (eg demographic, clinical, social) and information on exposures and potential confounders  **Addressed. Participant demographics, swab type, viral detection, and other relevant variables are reported in Table 1 and Supplementary Tables S4–S5.** | 8 |
|  |  | (b) Indicate number of participants with missing data for each variable of interest  **Supplementary Tables S4–S5 indicate availability of reliable microbiome profiles and report NA values where variables were not applicable.** |  |
|  |  | (c) Summarise follow-up time (eg, average and total amount)  **The manuscript provides ages at sampling, median intervals between paired asymptomatic and symptomatic swabs, and first-year outcomes.** |  |
| Outcome data | 15* | Report numbers of outcome events or summary measures over time  **Addressed. The manuscript reports wheezing episodes, bronchiolitis, symptomatic swab counts, age at first wheeze, and related summary measures.** | 10-12 |

| Main results | 16 | (*a*) Give unadjusted estimates and, if applicable, confounder-adjusted estimates and their precision (eg, 95% confidence interval). Make clear which confounders were adjusted for and why they were included  **Addressed. Regression estimates, IRRs, ORs, β coefficients, p-values, and 95% confidence intervals are reported, including models adjusted for sex where relevant.** | 10-12 |
| --- | --- | --- | --- |
|  |  | (*b*) Report category boundaries when continuous variables were categorized  **Continuous variables were not extensively categorized into arbitrary cut-points; where grouping was used, it mainly involved model-derived endotypes or predefined sampling categories. This item has limited applicability here.** |  |
|  |  | (*c*) If relevant, consider translating estimates of relative risk into absolute risk for a meaningful time period  **Results are reported mainly as IRRs, ORs, and regression estimates.** |  |
| Other analyses | 17 | Report other analyses done—eg analyses of subgroups and interactions, and sensitivity analyses  **Addressed. The manuscript reports subgroup analyses, interaction modelling, endotype transition analyses, and sensitivity analyses.** | 11-12 |
| Discussion | | | |
| Key results | 18 | Summarise key results with reference to study objectives  **Addressed in the opening paragraphs of the Discussion and Conclusion.** | 12-13 |
| Limitations | 19 | Discuss limitations of the study, taking into account sources of potential bias or imprecision. Discuss both direction and magnitude of any potential bias  **Addressed. Limitations include pilot sample size, single-centre design, altered viral circulation during the COVID period, sensitivity to outliers, limited longitudinal depth, and the constraints of 16S amplicon sequencing.** | 16 |
| Interpretation | 20 | Give a cautious overall interpretation of results considering objectives, limitations, multiplicity of analyses, results from similar studies, and other relevant evidence  **Addressed. The manuscript provides a balanced interpretation and explicitly notes the exploratory nature of the findings and the need for validation in larger cohorts.** | 13-16 |
| Generalisability | 21 | Discuss the generalisability (external validity) of the study results  **Addressed. External validity concerns are discussed, particularly the pilot, single-centre nature of the study.** | 16 |
| Other information | | | |
| Funding | 22 | Give the source of funding and the role of the funders for the present study and, if applicable, for the original study on which the present article is based  **Acknowledgments and funding section (p. 18). Role of funders not influencing study conduct is implied.** | 18 |

*Give information separately for exposed and unexposed groups.

**Note:** An Explanation and Elaboration article discusses each checklist item and gives methodological background and published examples of transparent reporting. The STROBE checklist is best used in conjunction with this article (freely available on the Web sites of PLoS Medicine at http://www.plosmedicine.org/, Annals of Internal Medicine at http://www.annals.org/, and Epidemiology at http://www.epidem.com/). Information on the STROBE Initiative is available at http://www.strobe-statement.org.
