## Supplemental Material for "Insights into the relationship between nasal bacterial composition and susceptibility to early-life respiratory disease: a pilot observational study"

**SUPPPLEMENTAL MATERIAL**

**Table S1.** Taxonomic path for the ASVs identified as potential contaminants by the R package decontam (1).

|  | Taxonomy (class; order; family; genus; specie) |
| --- | --- |
| Contaminant 1 | Bdellovibrionia_A;CAIPTA01;CAIPTA01;CAIPTA01;CAIPTA01 sp903867855 |
| Contaminant 2 | Gammaproteobacteria;Burkholderiales;Burkholderiaceae;Pelomonas;Pelomonas saccharophila |
| Contaminant 3 | Gammaproteobacteria;Burkholderiales;Burkholderiaceae;Variovorax;Variovorax sp003952185 |
| Contaminant 4 | Gammaproteobacteria;Nevskiales;Nevskiaceae;Nevskia;Nevskia soli |
| Contaminant 5 | Gammaproteobacteria;Pseudomonadales;Pseudomonadaceae;Pseudomonas_E; |
| Contaminant 6 | Gammaproteobacteria;Pseudomonadales;Pseudomonadaceae;Pseudomonas_E;Pseudomonas_E edaphica |
| Contaminant 7 | Gammaproteobacteria;Pseudomonadales;Pseudomonadaceae;Pseudomonas_E;Pseudomonas_E helleri |
| Contaminant 8 | Gammaproteobacteria;Steroidobacterales;Steroidobacteraceae;ZC4RG30;ZC4RG30 sp016720075 |

**Table S2.** Absolute read counts for the ASVs identified as potential contaminants by the R package decontam (1). Swabs specimens are identified by an alphanumeric code. The last three rows represent negative extraction controls.

| Sample | Contaminant 1 | Contaminant 2 | Contaminant 3 | Contaminant 4 | Contaminant 5 | Contaminant 6 | Contaminant 7 | Contaminant 8 |
| --- | --- | --- | --- | --- | --- | --- | --- | --- |
| MIC001 | 0 | 0 | 0 | 0 | 0 | 0 | 0 | 0 |
| MIC002 | 0 | 0 | 0 | 0 | 0 | 0 | 0 | 0 |
| MIC004 | 0 | 24 | 0 | 63 | 0 | 23 | 0 | 57 |
| MIC006 | 0 | 0 | 0 | 0 | 0 | 0 | 0 | 0 |
| MIC007 | 0 | 75 | 0 | 0 | 0 | 0 | 0 | 0 |
| MIC008 | 41 | 0 | 0 | 0 | 0 | 0 | 0 | 0 |
| MIC009 | 0 | 0 | 0 | 0 | 0 | 0 | 0 | 0 |
| MIC010 | 0 | 0 | 0 | 0 | 0 | 0 | 0 | 0 |
| MIC011 | 0 | 0 | 0 | 0 | 0 | 0 | 0 | 0 |
| MIC012 | 8 | 0 | 0 | 0 | 0 | 0 | 0 | 0 |
| MIC014 | 0 | 0 | 0 | 0 | 0 | 0 | 0 | 0 |
| MIC015 | 0 | 0 | 0 | 0 | 0 | 0 | 0 | 0 |
| MIC016 | 0 | 0 | 0 | 0 | 0 | 0 | 0 | 0 |
| MIC017 | 0 | 0 | 0 | 0 | 0 | 0 | 0 | 0 |
| MIC018 | 0 | 0 | 0 | 0 | 0 | 0 | 0 | 0 |
| MIC019 | 0 | 0 | 0 | 0 | 0 | 0 | 0 | 0 |
| MIC022 | 0 | 0 | 0 | 0 | 0 | 0 | 0 | 0 |
| MIC025 | 30 | 0 | 0 | 0 | 0 | 49 | 47 | 82 |
| MIC027 | 0 | 0 | 0 | 0 | 0 | 0 | 0 | 0 |
| MIC028 | 0 | 0 | 0 | 0 | 0 | 0 | 0 | 0 |
| MIC029 | 0 | 0 | 0 | 0 | 0 | 0 | 0 | 0 |
| MIC032 | 0 | 0 | 0 | 0 | 0 | 0 | 0 | 0 |
| MIC033 | 0 | 0 | 0 | 0 | 0 | 0 | 0 | 0 |
| MIC034 | 0 | 0 | 0 | 0 | 0 | 9 | 0 | 0 |
| MIC035 | 0 | 0 | 0 | 0 | 0 | 0 | 0 | 0 |
| MIC036 | 0 | 0 | 0 | 0 | 0 | 0 | 0 | 0 |
| MIC037 | 0 | 0 | 0 | 0 | 0 | 0 | 0 | 0 |
| MIC038 | 0 | 0 | 0 | 0 | 0 | 0 | 0 | 0 |
| MIC041 | 0 | 0 | 0 | 0 | 0 | 0 | 0 | 0 |
| MIC042 | 23 | 26 | 0 | 0 | 51 | 63 | 43 | 0 |
| MIC043 | 0 | 0 | 0 | 0 | 0 | 0 | 0 | 0 |
| MIC044 | 0 | 0 | 0 | 0 | 0 | 0 | 0 | 0 |
| MIC045 | 0 | 0 | 0 | 0 | 0 | 0 | 0 | 0 |
| MIC046 | 0 | 0 | 0 | 0 | 0 | 0 | 0 | 0 |
| MIC047 | 0 | 0 | 0 | 0 | 0 | 0 | 0 | 0 |
| MIC048 | 0 | 0 | 0 | 0 | 0 | 0 | 0 | 0 |
| MIC050 | 0 | 0 | 0 | 0 | 0 | 0 | 0 | 0 |
| MIC051 | 0 | 0 | 0 | 0 | 0 | 0 | 0 | 0 |
| MIC052 | 0 | 0 | 0 | 0 | 0 | 0 | 0 | 0 |
| MIC053 | 0 | 0 | 124 | 0 | 0 | 0 | 0 | 0 |
| MIC054 | 0 | 0 | 0 | 0 | 0 | 0 | 0 | 0 |
| MIC055 | 0 | 0 | 0 | 0 | 0 | 0 | 0 | 0 |
| MIC056 | 0 | 0 | 0 | 0 | 0 | 0 | 0 | 0 |
| MIC058 | 0 | 0 | 0 | 0 | 0 | 0 | 0 | 0 |
| MIC059 | 13 | 0 | 0 | 0 | 0 | 0 | 0 | 0 |
| MIC060 | 0 | 101 | 0 | 0 | 0 | 54 | 0 | 103 |
| MIC061 | 0 | 0 | 0 | 0 | 0 | 0 | 0 | 0 |
| MIC062 | 0 | 0 | 0 | 0 | 0 | 0 | 0 | 0 |
| MIC063 | 0 | 0 | 0 | 0 | 0 | 0 | 0 | 0 |
| MIC064 | 0 | 0 | 0 | 0 | 0 | 0 | 0 | 0 |
| MIC066 | 0 | 0 | 0 | 0 | 0 | 0 | 223 | 0 |
| MIC067 | 91 | 0 | 0 | 0 | 0 | 57 | 0 | 143 |
| MIC068 | 0 | 0 | 0 | 0 | 0 | 0 | 0 | 0 |
| MIC069 | 5 | 66 | 0 | 0 | 0 | 21 | 0 | 0 |
| MIC071 | 0 | 0 | 0 | 0 | 0 | 0 | 0 | 0 |
| MIC072 | 44 | 0 | 81 | 0 | 14 | 0 | 0 | 80 |
| MIC073 | 2 | 0 | 0 | 0 | 0 | 0 | 0 | 10 |
| MIC074 | 27 | 0 | 0 | 0 | 0 | 0 | 0 | 0 |
| MIC075 | 33 | 12 | 0 | 0 | 13 | 29 | 31 | 29 |
| MIC076 | 0 | 0 | 0 | 0 | 0 | 0 | 0 | 0 |
| MIC077 | 0 | 0 | 0 | 0 | 0 | 0 | 0 | 0 |
| MIC078 | 0 | 0 | 0 | 0 | 0 | 0 | 0 | 0 |
| MIC080 | 0 | 162 | 77 | 73 | 0 | 0 | 0 | 0 |
| MIC081 | 16 | 18 | 0 | 0 | 0 | 0 | 0 | 0 |
| MIC082 | 149 | 19 | 0 | 0 | 0 | 0 | 0 | 95 |
| MIC083 | 0 | 0 | 0 | 0 | 0 | 0 | 0 | 0 |
| MIC084 | 0 | 23 | 0 | 0 | 0 | 0 | 4 | 0 |
| MIC085 | 24 | 8 | 0 | 0 | 0 | 0 | 0 | 22 |
| MIC087 | 0 | 0 | 0 | 0 | 0 | 0 | 0 | 0 |
| MIC088 | 0 | 0 | 0 | 0 | 0 | 0 | 0 | 0 |
| MIC089 | 0 | 0 | 0 | 0 | 0 | 0 | 0 | 0 |
| MIC090 | 0 | 83 | 0 | 0 | 0 | 0 | 0 | 54 |
| NEG_B3 | 97 | 96 | 40 | 10 | 29 | 27 | 24 | 17 |
| NEG1_B1 | 19 | 120 | 0 | 0 | 32 | 0 | 0 | 28 |
| NEG2_B1 | 164 | 232 | 0 | 113 | 24 | 188 | 126 | 111 |

**Table S3.** Outcomes of the permutational test of significance of the Procrustes results showed in Figure S3. The results show a strong correlation coefficient and a small value for the goodness-of-fit statistic M2. These results indicate a substantial agreement between the compared datasets and suggest that the filtering steps had minimal impact on the structure of the original dataset (Raw counts).

| Datasets | Procrustes Sum of Squares (M^2^) | Correlation in a symmetric Procrustes rotation | Significance |
| --- | --- | --- | --- |
| Raw counts Vs Removed low counts/singletons/Unclassified and contaminants | 0.002 | 0.99 | 0.00001 |

**Table S4.** Metadata for asymptomatic nasal swabs included in this study. For each infant, columns indicate whether the swab yielded a reliable microbiome profile (Nasal microbiota, Yes = included in analyses; No = excluded as no reliable microbial profile), season of collection, sex, virus detected, childcare attendance (ever), and clinical data related to respiratory phenotypes: ever bronchiolitis (Ever bronc), ever wheeze (Ever whe), ever recurrent wheeze (Ever rec whe), number of wheezing episodes in the first year of life (# whe epi), and number of symptomatic swabs in the first year of life (# symp swabs). Abbreviations: #, number; Asy, asymptomatic; S-S, summer-spring; W-A, winter-autumn; F, female; M, male; SARS-Co-2, Severe acute respiratory syndrome coronavirus 2; RHV, rhinovirus; cc, childcare; bronc, bronchiolitis; whe, wheeze; rec whe, recurrent wheeze. For one infant (infant 15), two asymptomatic swabs were present in this cohort, but only one swab yielded microbiome data.

| Study ID | Nasal microbiota | Infant # | Swab  type | Season at collection | Sex | Virus detected | Ever cc | Ever bronc | Ever whe | Ever rec whe | # whe epi | # symp swabs |
| --- | --- | --- | --- | --- | --- | --- | --- | --- | --- | --- | --- | --- |
| MIC008 | Yes | Infant_7 | Asy | S-S | F | negative | Yes | No | No | No | 0 | 3 |
| MIC010 | Yes | Infant_8 | Asy | S-S | M | negative | Yes | Yes | Yes | No | 1 | 3 |
| MIC012 | Yes | Infant_9 | Asy | W-A | M | negative | No | Yes | Yes | Yes | 10 | 4 |
| MIC014 | Yes | Infant_10 | Asy | W-A | M | negative | Yes | No | Yes | No | 1 | 1 |
| MIC016 | Yes | Infant_11 | Asy | W-A | F | negative | No | Yes | Yes | Yes | 14 | 3 |
| MIC018 | Yes | Infant_12 | Asy | S-S | M | negative | Yes | No | No | No | 0 | 5 |
| MIC021 | No | Infant_14 | Asy | W-A | M | negative | Yes | No | No | No | 0 | 3 |
| MIC024 | No | Infant_15 | Asy | S-S | M | negative | No | No | No | No | 0 | 5 |
| MIC025 | Yes | Infant_15 | Asy | W-A | M | negative | No | No | No | No | 0 | 5 |
| MIC027 | Yes | Infant_16 | Asy | S-S | M | negative | Yes | No | Yes | Yes | 2 | 3 |
| MIC029 | Yes | Infant_17 | Asy | W-A | M | negative | Yes | Yes | Yes | No | 1 | 4 |
| MIC031 | No | Infant_18 | Asy | S-S | F | negative | No | Yes | No | No | 0 | 1 |
| MIC033 | Yes | Infant_19 | Asy | W-A | M | negative | No | No | Yes | No | 1 | 6 |
| MIC034 | Yes | Infant_20 | Asy | W-A | M | negative | No | No | No | No | 0 | 4 |
| MIC035 | Yes | Infant_21 | Asy | W-A | F | negative | No | No | Yes | No | 1 | 4 |
| MIC037 | Yes | Infant_22 | Asy | W-A | F | negative | Yes | No | Yes | Yes | 2 | 5 |
| MIC040 | No | Infant_24 | Asy | S-S | F | negative | Yes | No | Yes | No | 1 | 4 |
| MIC042 | Yes | Infant_25 | Asy | W-A | F | negative | No | No | No | No | 0 | 4 |
| MIC044 | Yes | Infant_26 | Asy | W-A | M | negative | No | No | No | No | 0 | 3 |
| MIC046 | Yes | Infant_27 | Asy | S-S | F | negative | No | Yes | No | No | 0 | 3 |
| MIC048 | Yes | Infant_28 | Asy | W-A | M | negative | Yes | No | Yes | Yes | 5 | 3 |
| MIC050 | Yes | Infant_29 | Asy | W-A | M | negative | Yes | No | No | No | 0 | 5 |
| MIC052 | Yes | Infant_30 | Asy | W-A | F | negative | No | No | Yes | No | 1 | 2 |
| MIC055 | Yes | Infant_32 | Asy | W-A | M | negative | Yes | No | No | No | 0 | 2 |
| MIC056 | Yes | Infant_33 | Asy | W-A | F | negative | Yes | Yes | Yes | No | 1 | 2 |
| MIC058 | Yes | Infant_34 | Asy | W-A | M | negative | No | No | Yes | Yes | 2 | 7 |
| MIC060 | Yes | Infant_35 | Asy | W-A | F | Negative | Yes | No | Yes | Yes | 2 | 6 |
| MIC062 | Yes | Infant_36 | Asy | W-A | F | negative | No | No | No | No | 0 | 2 |
| MIC064 | Yes | Infant_37 | Asy | W-A | F | RHV | Yes | No | Yes | Yes | 4 | 2 |
| MIC065 | No | Infant_38 | Asy | W-A | F | negative | No | No | No | No | 0 | 1 |
| MIC066 | Yes | Infant_39 | Asy | W-A | F | negative | No | No | No | No | 0 | 4 |
| MIC067 | Yes | Infant_40 | Asy | W-A | F | negative | No | Yes | Yes | No | 1 | 2 |
| MIC069 | Yes | Infant_42 | Asy | W-A | M | negative | Yes | No | Yes | Yes | 6 | 1 |
| MIC071 | Yes | Infant_43 | Asy | W-A | M | negative | No | No | No | No | 0 | 3 |
| MIC073 | Yes | Infant_44 | Asy | W-A | F | negative | No | No | Yes | No | 1 | 2 |
| MIC075 | Yes | Infant_45 | Asy | W-A | M | negative | Yes | Yes | No | No | 0 | 4 |
| MIC076 | Yes | Infant_46 | Asy | W-A | M | negative | Yes | No | Yes | Yes | 2 | 7 |
| MIC078 | Yes | Infant_47 | Asy | W-A | F | negative | No | No | Yes | No | 1 | 6 |
| MIC082 | Yes | Infant_50 | Asy | W-A | F | negative | No | No | Yes | No | 1 | 1 |
| MIC085 | Yes | Infant_52 | Asy | S-S | F | negative | No | No | Yes | No | 1 | 2 |
| MIC087 | Yes | Infant_53 | Asy | S-S | M | SARS-CoV-2 | No | No | Yes | Yes | 6 | 4 |
| MIC090 | Yes | Infant_55 | Asy | S-S | M | negative | No | No | No | No | 0 | 1 |

**Table S5.** Metadata for symptomatic nasal swabs included in this study. For each infant, columns indicate whether the swab yielded a reliable microbiome profile (Nasal microbiota, Yes = included in analyses; No = excluded because it did not yield a reliable microbial profile), season of collection, sex, virus detected, whether an asymptomatic swab for this infant is available (Paired, Yes), and the difference in days between the date of sampling of the symptomatic and asymptomatic swab (Difference with asymptomatic). NA indicates not applicable (not observed); for example, difference with asymptomatic is NA when Paired = No. Abbreviations: #, number; Sym, symptomatic; S-S, summer-spring; W-A, winter-autumn; F, female; M, male; SARS-Co-2, severe acute respiratory syndrome coronavirus 2; RHV, rhinovirus; RSV, respiratory syncytial virus; HMPV, human metapneumovirus. For one infant (infant 15), two symptomatic swabs were present in this cohort, but only one swab yielded microbiome data.

| Study ID | Nasal microbiota | Infant # | Swab  type | Season at collection | Sex | Virus detected | Paired | Difference with asymptomatic (days) |
| --- | --- | --- | --- | --- | --- | --- | --- | --- |
| MIC001 | Yes | Infant_1 | Sym | S-S | F | SARS-CoV-2 | No | NA |
| MIC002 | Yes | Infant_2 | Sym | W-A | F | SARS-CoV-2 | No | NA |
| MIC003 | No | Infant_3 | Sym | S-S | F | SARS-CoV-2, RHV, Adenovirus | No | NA |
| MIC004 | Yes | Infant_4 | Sym | W-A | F | SARS-CoV-2 | No | NA |
| MIC005 | No | Infant_5 | Sym | S-S | F | RHV | No | NA |
| MIC006 | Yes | Infant_6 | Sym | S-S | F | SARS-CoV-2, RHV | No | NA |
| MIC007 | Yes | Infant_7 | Sym | S-S | F | RHV | Yes | 108 |
| MIC009 | Yes | Infant_8 | Sym | S-S | M | RSV | Yes | 42 |
| MIC011 | Yes | Infant_9 | Sym | W-A | M | RHV | Yes | 27 |
| MIC013 | No | Infant_10 | Sym | W-A | M | RHV | Yes | 62 |
| MIC015 | Yes | Infant_11 | Sym | W-A | F | RHV | Yes | 19 |
| MIC017 | Yes | Infant_12 | Sym | W-A | M | SARS-CoV-2, RHV | Yes | 161 |
| MIC019 | Yes | Infant_13 | Sym | W-A | M | SARS-CoV-2, RHV | No | NA |
| MIC020 | No | Infant_14 | Sym | W-A | M | Parainfluenza, RHV, Adenovirus | Yes | 28 |
| MIC022 | Yes | Infant_15 | Sym | W-A | M | Parainfluenza | Yes | 12 |
| MIC023 | No | Infant_15 | Sym | W-A | M | SARS-CoV-2 | Yes | 78 |
| MIC026 | No | Infant_16 | Sym | S-S | M | RHV | Yes | 61 |
| MIC028 | Yes | Infant_17 | Sym | W-A | M | RHV | Yes | 32 |
| MIC030 | No | Infant_18 | Sym | W-A | F | SARS-CoV-2 | Yes | 88 |
| MIC032 | Yes | Infant_19 | Sym | W-A | M | RHV | Yes | 25 |
| MIC036 | Yes | Infant_22 | Sym | W-A | F | RHV | Yes | 41 |
| MIC038 | Yes | Infant_23 | Sym | W-A | F | SARS-CoV-2 | No | NA |
| MIC039 | No | Infant_24 | Sym | W-A | F | RHV | Yes | 105 |
| MIC041 | No | Infant_25 | Sym | W-A | F | RHV | Yes | 44 |
| MIC043 | Yes | Infant_26 | Sym | W-A | M | RHV | Yes | 61 |
| MIC045 | Yes | Infant_27 | Sym | W-A | F | SARS-CoV-2 | Yes | 63 |
| MIC047 | Yes | Infant_28 | Sym | W-A | M | SARS-CoV-2 | Yes | 40 |
| MIC049 | No | Infant_29 | Sym | W-A | M | RHV | Yes | 61 |
| MIC051 | Yes | Infant_30 | Sym | S-S | F | RHV | Yes | 35 |
| MIC053 | Yes | Infant_31 | Sym | W-A | M | SARS-CoV-2 | No | NA |
| MIC054 | Yes | Infant_32 | Sym | W-A | M | RHV | Yes | 33 |
| MIC057 | No | Infant_34 | Sym | W-A | M | RHV | Yes | 29 |
| MIC059 | Yes | Infant_35 | Sym | W-A | F | RHV | Yes | 20 |
| MIC061 | Yes | Infant_36 | Sym | W-A | F | RHV | Yes | 73 |
| MIC063 | Yes | Infant_37 | Sym | S-S | F | SARS-CoV-2 | Yes | 55 |
| MIC068 | Yes | Infant_41 | Sym | W-A | F | SARS-CoV-2 | No | NA |
| MIC070 | No | Infant_43 | Sym | S-S | M | RHV | Yes | 18 |
| MIC072 | Yes | Infant_44 | Sym | W-A | F | RHV | Yes | 61 |
| MIC074 | Yes | Infant_45 | Sym | W-A | M | RHV | Yes | 7 |
| MIC077 | Yes | Infant_47 | Sym | W-A | F | RHV | Yes | 9 |
| MIC079 | No | Infant_48 | Sym | W-A | M | SARS-CoV-2 | No | NA |
| MIC080 | Yes | Infant_49 | Sym | W-A | F | SARS-CoV-2 | No | NA |
| MIC081 | Yes | Infant_50 | Sym | W-A | F | HMPV | Yes | 67 |
| MIC083 | Yes | Infant_51 | Sym | W-A | M | SARS-CoV-2 | No | NA |
| MIC084 | Yes | Infant_52 | Sym | S-S | F | SARS-CoV-2 | Yes | 34 |
| MIC086 | No | Infant_53 | Sym | S-S | M | SARS-CoV-2 | Yes | 1 |
| MIC088 | Yes | Infant_54 | Sym | W-A | F | SARS-CoV-2 | No | NA |
| MIC089 | Yes | Infant_55 | Sym | S-S | M | SARS-CoV-2 | Yes | 33 |

**Supplemental Material and Methods**

**Symptomatic swab definition**

Symptomatic swabs were defined by either a temperature >37.5°C along with at least one symptom (runny/blocked nose, cough, sneeze, headache, myalgia, chills, rigors, tiredness, sore throat, dyspnea, loss of taste/smell, poor feeding, or diarrhea), or a temperature <37.5°C with at least two respiratory symptoms present (2).

**Sample collection**

Both nostrils were sampled at scheduled visits and when a symptomatic episode was recorded through the smartphone app. One swab (FLoQSwab®, Copan Group, Italy) was placed in viral transport media (eSwab, Copan Group, Italy) and assayed against a qPCR panel for 8 common respiratory viruses at commercial laboratory Western Diagnostics Pathology Pty Ltd, Western Australia (2). The qPCR panel was designed to detect Influenza “A” and “B”, respiratory syncytial virus, human metapneumovirus, parainfluenza, rhinovirus, adenovirus and SARS-CoV-2. The second swab was placed in 1 mL of eNAT® buffer (eNAT®, Copan Group, Italy), transported to the laboratory at 4ºC and subsequently stored at -80ºC until processing for microbiome analyses (2).

**Isolation of microbial DNA from nasal swabs, amplification of the full-length 16S rRNA gene and sequencing of the amplicon pools**

We extracted bacterial DNA from 400 µL of eNAT® suspension using the QIAamp DNA kit (QIAGEN) blinded to clinical data. We followed the manufacturer’s protocol and included two modifications: i) inclusion of a bead-beating pre-processing step using 0.1 mm diameter zirconia/silica beads (BioSpec) to enhance lysis efficiency and minimize extraction-related bias in community profiles, as we previously described (3), and ii) addition of 100 µg of yeast tRNA carrier (ThermoFisher) prior to the binding step to improve DNA recovery. To identify reagent-related contaminants, negative extraction controls were processed alongside the swab specimens. We used a full-length *16S rRNA* gene amplicon sequencing approach to profile the bacterial component of the nasal epithelium-associated microbiota. Amplicon library preparation and sequencing were done at the Australian Genome Research Facility (AGRF Ltd, Melbourne, VIC, Australia). Briefly, for amplification of the full-length *16S rRNA* bacterial gene we used the degenerated forward 5’-*GCATC/barcode/AGRGTTYGATYMTGGCTCAG* and reverse 5’-*GCATC/barcode/RGYTACCTTGTTACGACTT* primers following the protocol recommended by PacBio (<https://www.pacb.com/wp-content/uploads/Procedure-checklist-Amplification-of-bacterial-full-length-16S-rRNA-gene-with-barcoded-primers.pdf>). Individual PCR reactions showing the amplification of a single PCR product of the expected size are represented in Figure S1. Libraries were prepared using the SMRTbell® preparation kit 3.0, and sequenced using the Sequel II binding kit 3.1, on a single SMRT cell 8M, using a PacBio Sequel IIe instrument.

To minimize bias associated with amplifying and sequencing the *16SrRNA* gene in DNA extracts obtained from low bacterial biomass samples we followed two strategies: parallel DNA extraction and sequencing from eNAT® buffer negative controls, and bacterial load quantification of the DNA extracts using a pan bacteria TaqMan® assay (4). Details on the TaqMan® assay are described in our previous publication (5).

**Processing of the sequencing data**

Data was processed using *DADA2* (v1.28.0) (6), as implemented in the nfcore pipeline ampliseq (v2.8.0) (https://nfco.re/ampliseq), using Nextflow (v25.04.2) (7). For taxonomic classification of the generated amplicon sequencing variants (ASVs), we used the naïve Bayesian classifier implemented in *DADA2* (8), and a pretrained Genome Taxonomy Database (GTDB; vR09-RS220) as reference library (9). To prevent bias from potential contaminants and distinguish a true biological signal from background noise, we followed the analytical workflow outlined in our previous work (10). Briefly, only bacterial DNA extracts with qPCR result values higher than 3 standard deviations from the mean result in negative extraction controls were retained for further analyses (Figure S2). Then, the taxonomic profiles in the negative extraction controls were used to identify and remove potential contaminants using the R package *decontam* (v1.20.0) (1) (Table S1-S2). We performed Procrustes analysis to confirm that the described filtering steps did not significantly impact the overall structure of the original, unfiltered taxonomic data table (Table S3 and Figure S3). After filtering, we removed a sample with only 136 reads from further analyses.

**Statistical analysis**

Analyses were done in R statistical software (v4.3.0) within the RStudio environment (v2023.03.0). The R code and the deidentified datasets required to reproduce the findings of this study are available in the accompanying GitHub repository: <https://github.com/jacapmar/MicrobiomeAERIAL>. We used vegan (v2.6-4) for conducting permutational multivariate analysis of variance (PERMANOVA) and Procrustes analyses, and to calculate ecological diversity indexes. PERMANOVA was performed on a beta diversity distance matrix constructed using Aitchison distances. For analyses including repeated measures (e.g. asymptomatic versus symptomatic comparisons across all swabs), permutations were restricted within infant (infant number) to account for within-subject dependence. Forsensitivity analyses assessing the effect of age at sampling on beta diversity, PERMANOVA, was performed on asymptomatic swabs only. Principal Component Analysis (PCA) was done using the functions contained in the MixOmics package (v6.24.0) (11). To address the compositional nature of the microbiome data, a centered log-ratio (CLR) transformation was applied to the full taxonomy table before performing correlation-based analyses, including PCA and Spearman’s rank correlation. For probabilistic modelling of microbial compositional profiles, we used Dirichlet Multinomial Mixtures (DMM) models as implemented in the R package DirichletMultinomial (v1.42.0) (12). Differential abundance analysis was performed using the Analysis of Compositions of Microbiomes with Bias Correction framework, as implemented in the *ancombc2* function from the ANCOMBC R package (v2.2.0) (13). According to the normality of the data and the hypotheses being tested, we also used Student’s t-test and Wilcoxon Rank Sum Test. When multiple hypotheses were tested simultaneously, the false discovery rate method was used to control the type I error rate. Categorical data in contingency tables were compared using the Fisher exact test. Linear models were employed using the R built-in function *lm*. Logistic regression was fitted using the *glm* function in R. Count data was initially modelled using Poisson regression as implemented in the R built-in function *glm*. To evaluate the validity of the Poisson distributional assumption (that the mean equals the variance), we applied the *dispersiontest* function from the AER package in R (v1.2-14). This score test evaluates overdispersion based on Pearson residuals and the residual deviance. When overdispersion was detected (i.e., variance significantly greater than the mean), models were refitted using a negative binomial regression, which accounts for extra-Poisson variation by including a dispersion parameter. Negative binomial regression was fitted using the *glm.nb* function of the R package MASS (v7.3-60). For wheezing episode count models, endotype was the primary predictor and sex was included as an adjustment term. Sex-by-endotype interaction terms were included where indicated. Models of age at first wheeze were fitted using linear regression without additional covariate adjustment. Binary respiratory outcomes were analysed using logistic regression with sex included where indicated. To assess whether sampling time could confound asymptomatic/baseline community patterns, we evaluated age at sampling in asymptomatic swabs in relation to endotype assignment (logistic regression), alpha diversity (Spearman correlation), and beta diversity (PERMANOVA). To account for within-infant dependence in analyses comparing asymptomatic and symptomatic swabs across the full dataset, we fitted linear mixed-effects models with infant as a random intercept using the nlme package (v3.1-168) in R. Incidence rate ratios (IRRs) with 95% confidence intervals were calculated from the coefficients of the GLM and GLM.nb models. Marginal effects were estimated using the *ggpredict* function from the ggeffects package (v2.2.1) in R. Where estimated marginal means were compared, contrasts were tested using Wald z-test and multiplicity was controlled using appropriate adjustements (e.g. Dunnett for comparison to a reference group as specified). We used the R packages tidyverse (v2.0.0) for data manipulation and ggplot2 (v3.4.2) for data visualization. Figures were created using Adobe Illustrator (v24.1). The cut-off for statistical significance was set at *p*<0.05.

**Supplemental Figures S1-S15**

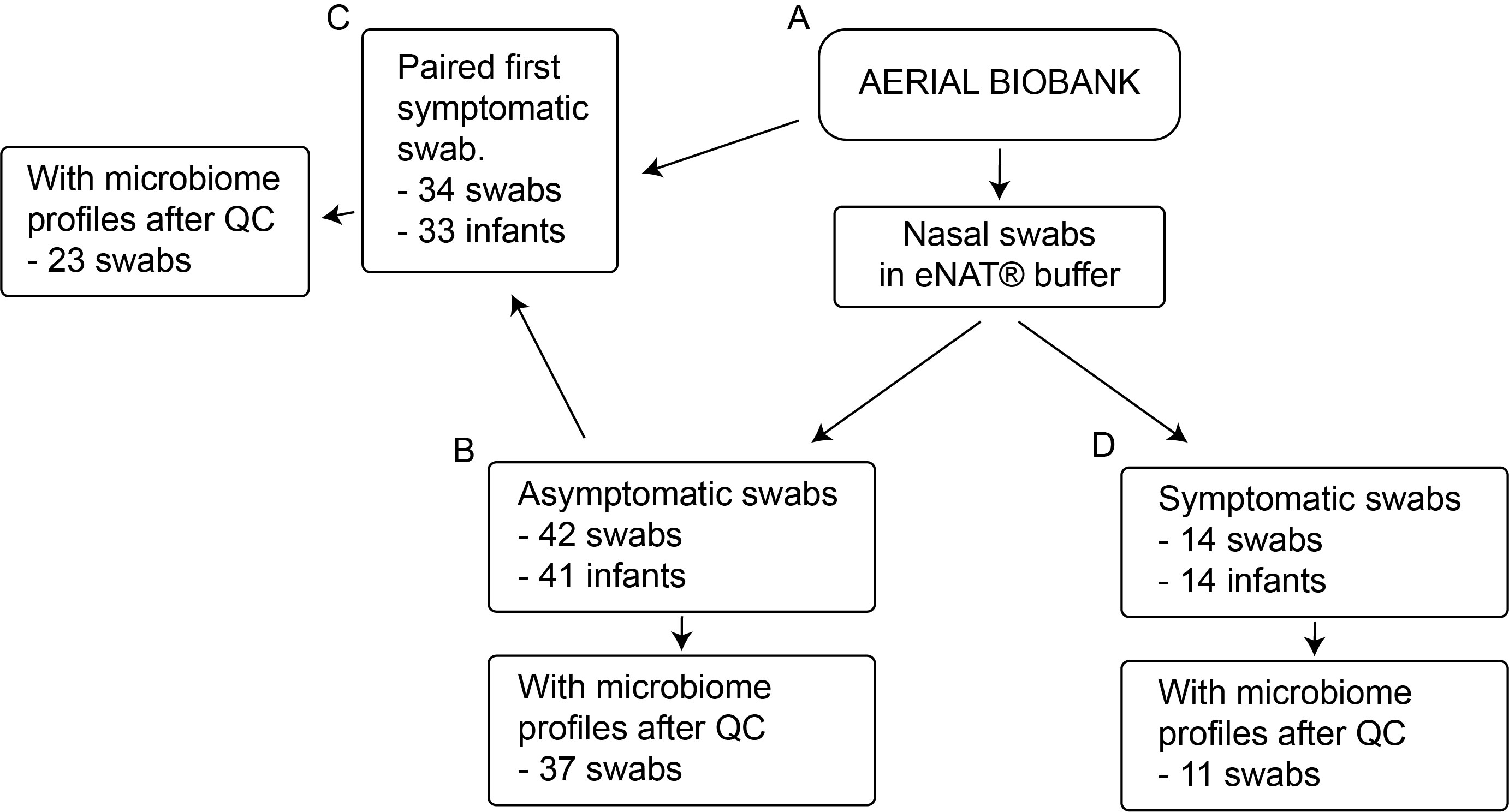

**Figure S1.** Flow diagram for nasal swab selection for this study. Schematic overview of the nasal swabs included in this pilot study. Nasal swabs in eNAT® buffer were obtained from the larger AERIAL cohort study (**A**). We randomly selected 42 asymptomatic swabs which represented 41 infants (**B**). For this subset of asymptomatic swabs, we selected an associated subsequent symptomatic episode (paired samples) for 34 swabs, representing 33 infants (**C**). We additionally included 14 symptomatic swabs without a preceding sequenced asymptomatic swab to complete the dataset (**D**). Next to each subset, we represent the number of swabs yielding usable microbiome profiles after sequencing quality control (QC). For one infant, two asymptomatic and two symptomatic swabs were present in this cohort, but only one asymptomatic and one symptomatic swab yielded microbiome data.

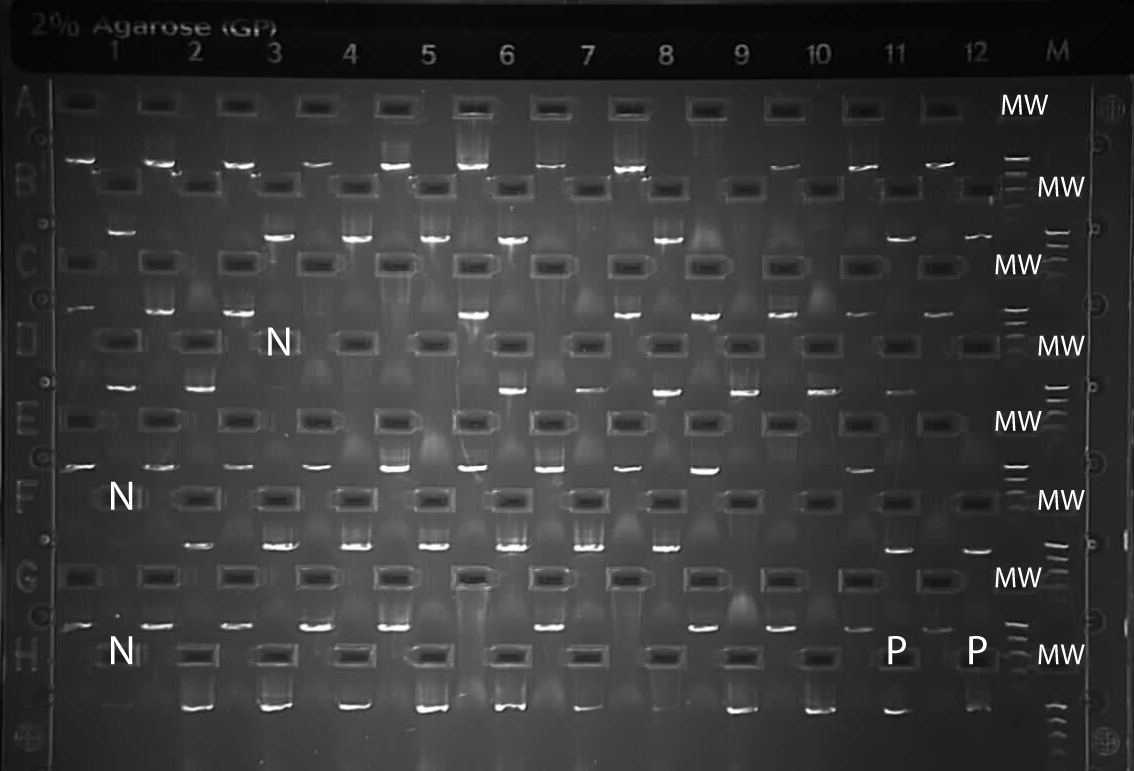

**Figure S2.** PCR-mediated amplification of the full length 16S rRNA gene in the microbial DNA extracts obtained in this study. PCR products were separated using a E-Gel™ 2% agarose gel (ThermoFisher), to confirm the amplification of a single specific product of the expected size (~1.5Kb). The gel lanes loaded with negative (N) and positive (P) extraction controls, or molecular weight reference (MW) are indicated.

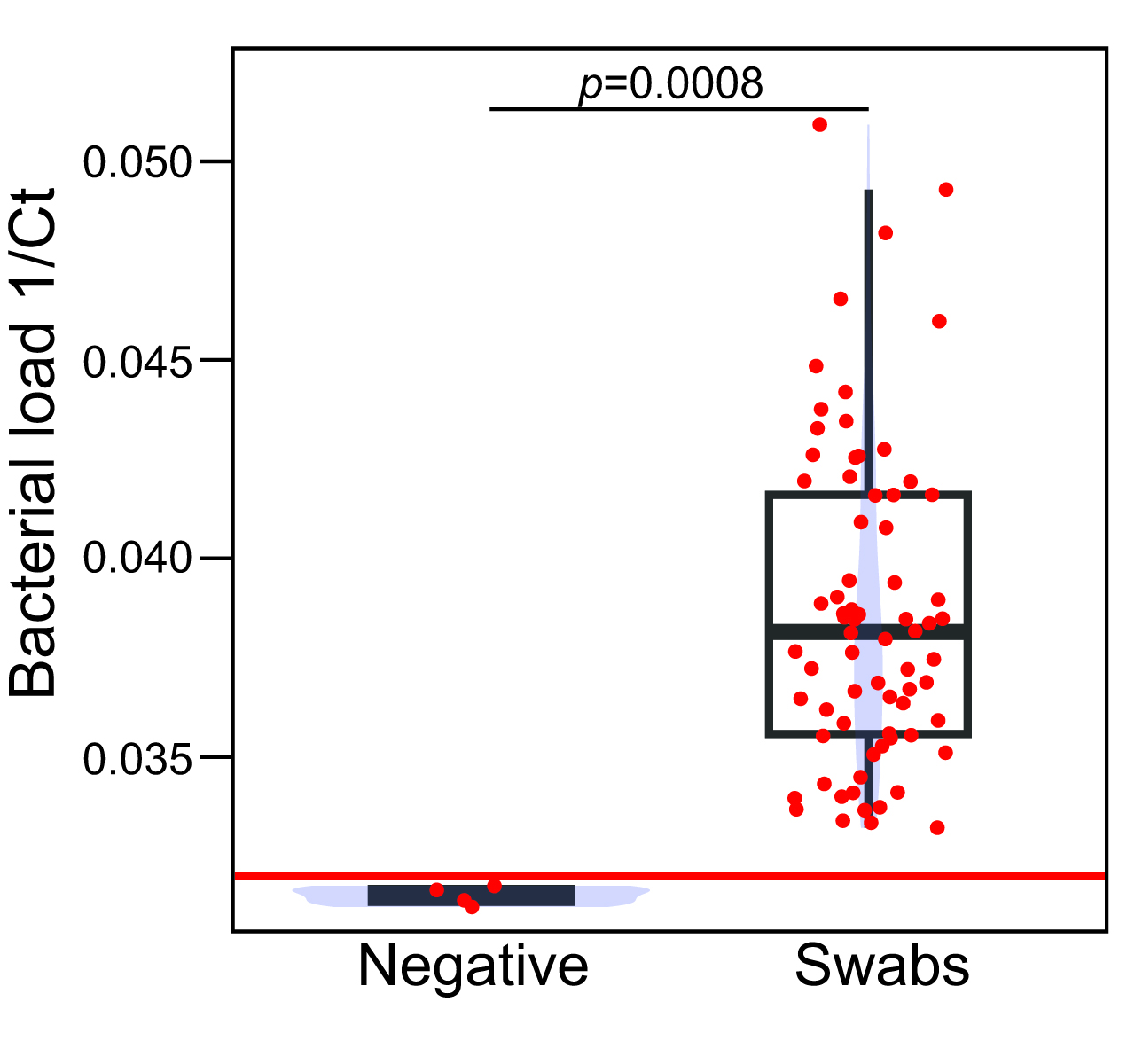

**Figure S3.** Quantification of bacterial load using the TaqMan® assay described in our previous publication (5). Individual data points with jitter are represented with red dots. The red line indicates the cut-off we set as lower limit of detection in our cohort, which lies 3 standard deviations from the mean bacterial load in the negative extraction controls.

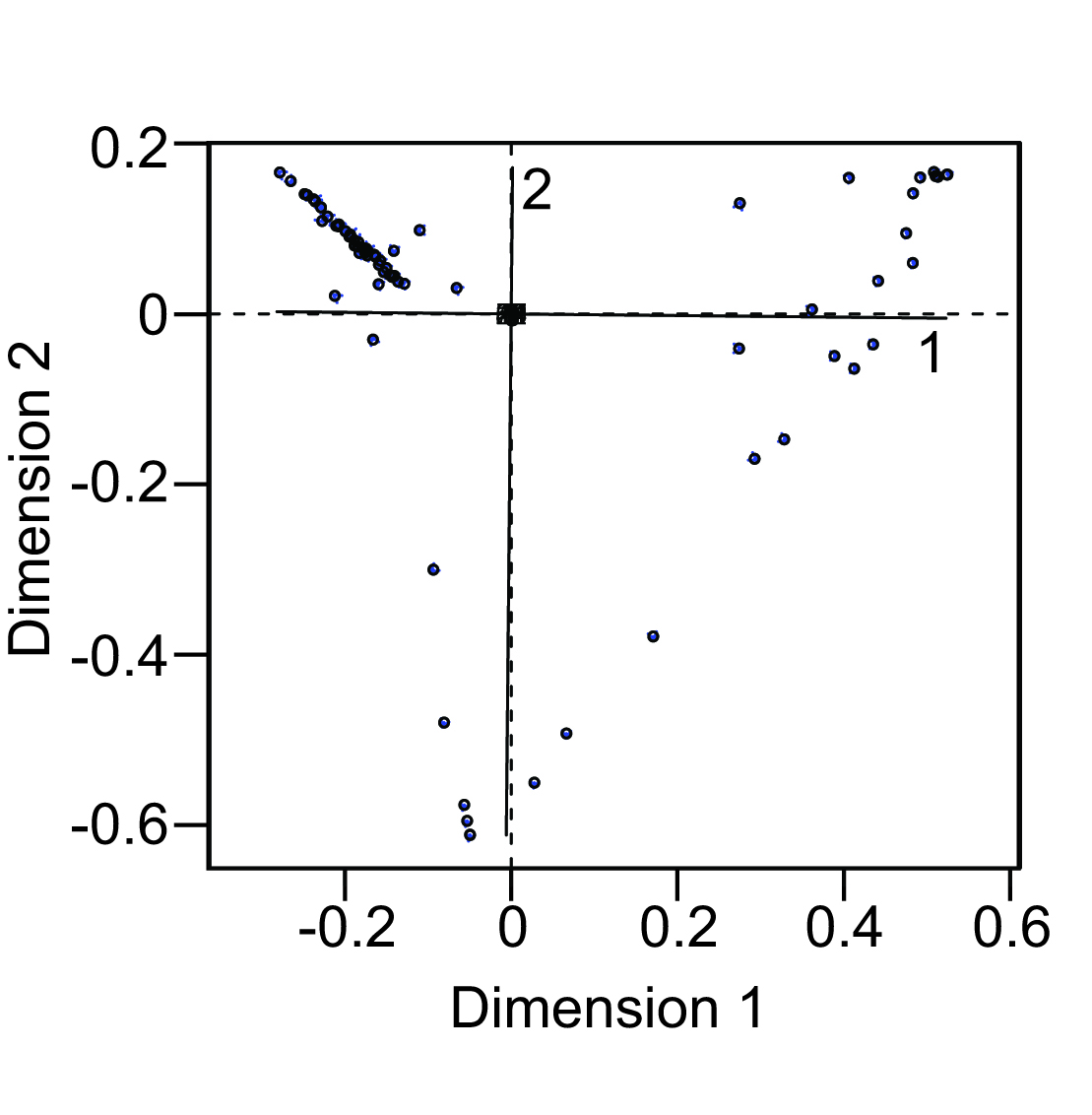

**Figure S4.** Procrustes analysis of the correlation between the unfiltered taxonomic data table, and the dataset after removing unclassified reads and potential contaminants identified using the R package *decontam* (1). The spatial location of the samples in the first ordination (filtered dataset) is indicated with circles. The location of the sample in the second ordination (unfiltered taxonomic table) is indicated with a blue arrow. Solid black lines indicate the rotation required to minimise the sum of squared distances between matching samples in both ordinations, while numbers indicate the axes to be rotated.

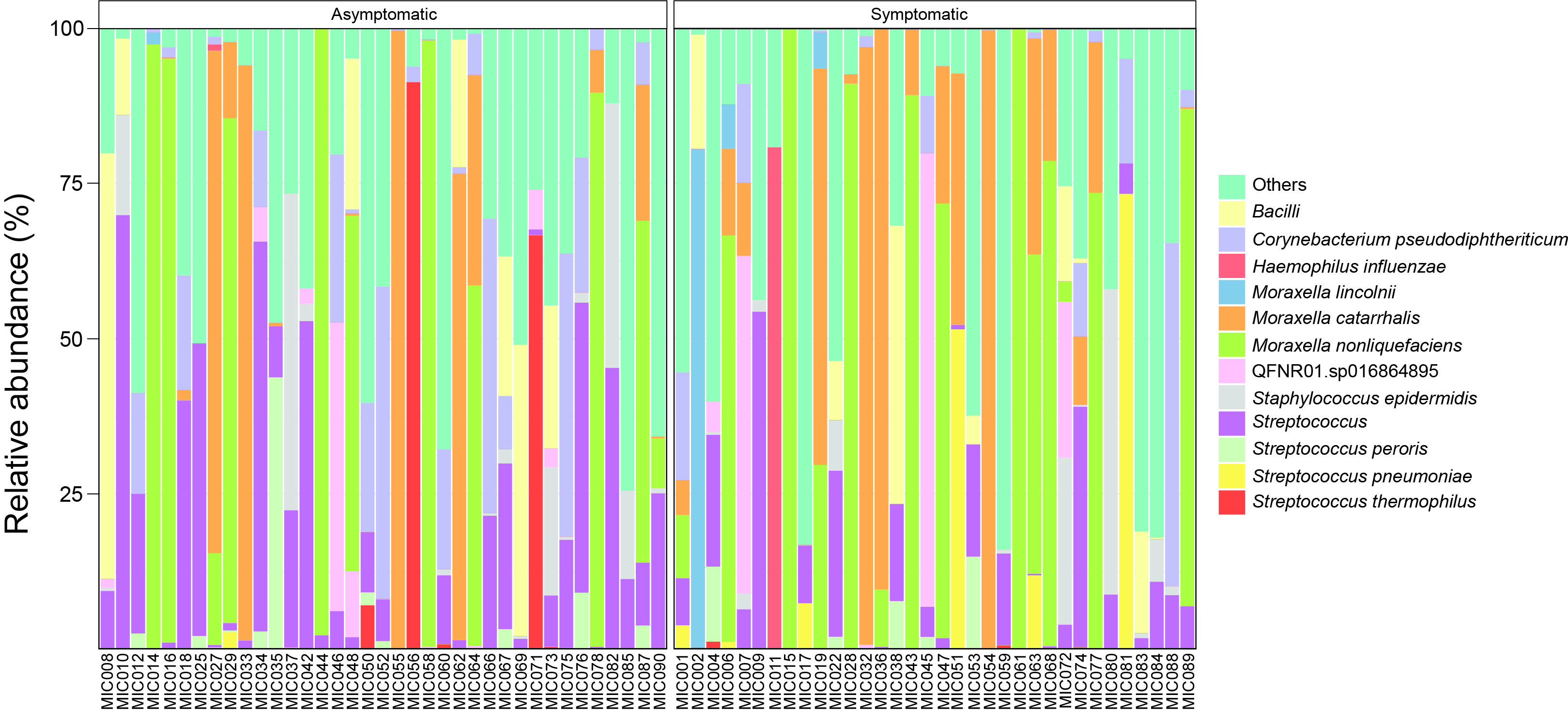

**Figure S5.** Bar plots representing the bacterial compositional profiles observed in the DNA extracts obtained from the nasal swabs included in this study. The top 11 amplicon sequencing variants (ASVs) and an ASV assigned to the *Haemophilus influenzae* taxon are represented. Profiles are grouped based on whether they represent the bacterial communities from swabs collected at scheduled visits (asymptomatic, left), or a profile associate with a symptomatic respiratory infection (symptomatic, right).

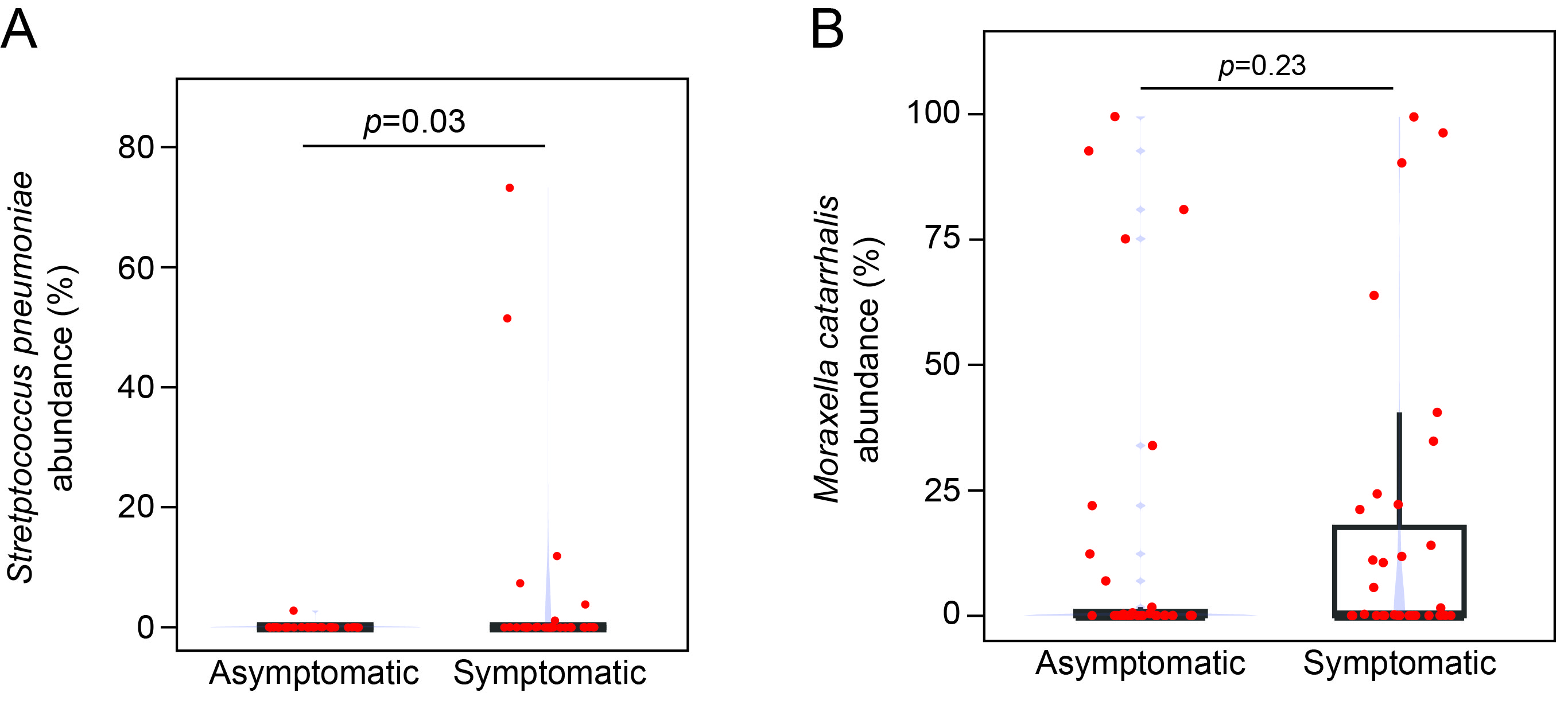

**Figure S6.** Barplots overlaid with density plots (blue) showing the abundance of *Streptococcus pneumoniae* (**A**) and *Moraxella catarrhalis* (**B**) in bacterial profiles from swabs collected during scheduled visits (asymptomatic) or during symptomatic episodes. Red dots represent individual samples. Groups were compared using the Wilcoxon rank-sum test (WRST), with p-values corrected for multiple comparisons using the false discovery rate (FDR) method, as shown in the corresponding barplots. *Streptococcus pneumoniae* was higher in symptomatic swabs (mean [SD]; asymptomatic, 0.1% [0.5] versus symptomatic, 4.4% [15.2]; WRST FDR-corrected (*r*=0.28, *p*=0.03)), whereas *Moraxella catarrhalis* did not differ (asymptomatic 11.5% [27.7] versus symptomatic, 16.1% [28.9]; WRST FDR-corrected (*r*=0.14, *p*=0.23))

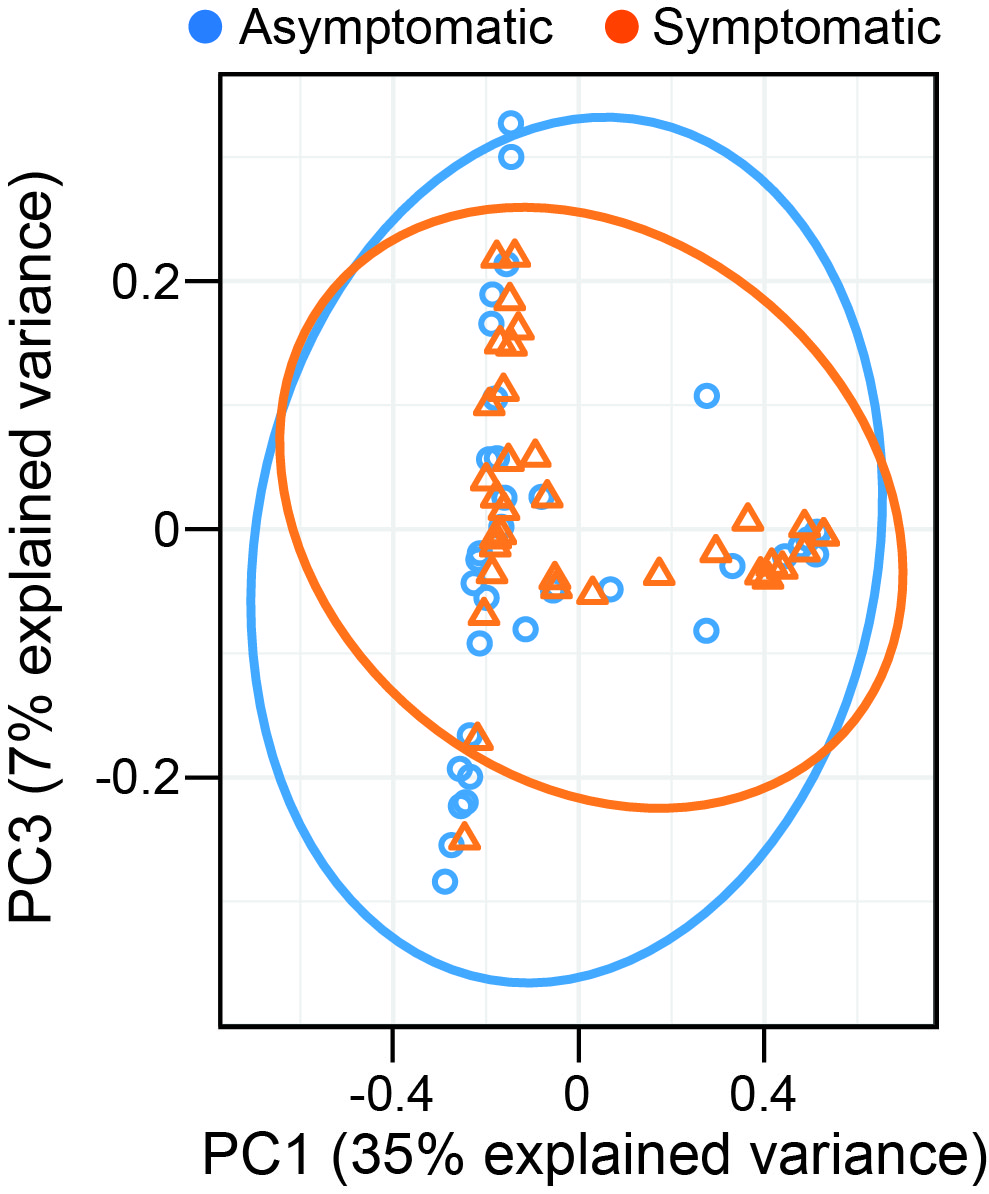

**Figure S7.** Principal component analysis (PCA) plot showing the linear projection of bacterial compositional profiles from each sample onto the components 1 and 3 of the PCA model. Dots represent individual bacterial profiles, coloured based on whether the sample is from an asymptomatic swab (asymptomatic, blue) or a symptomatic swab (symptomatic, orange).

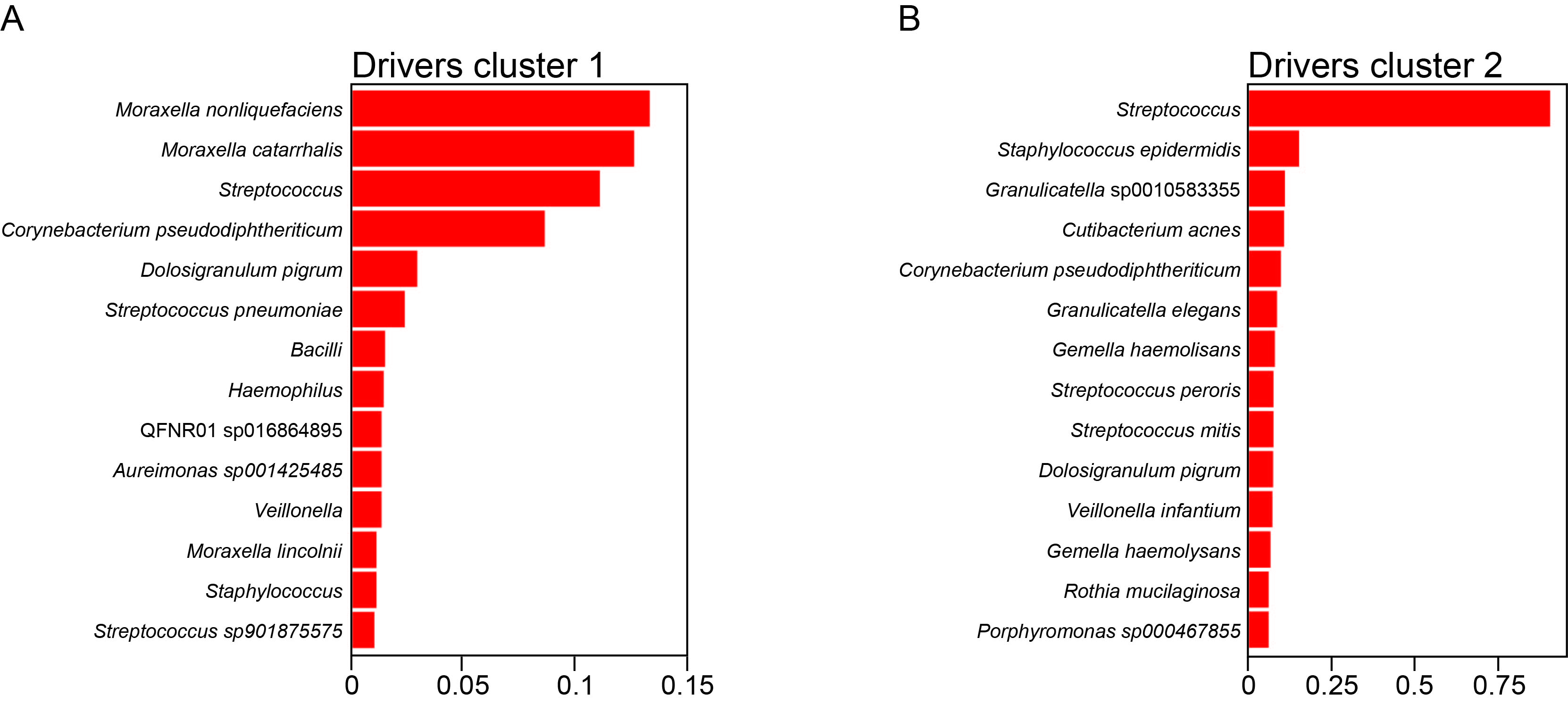

**Figure S8.** Bar plots represent the contribution of the indicated ASVs to each of the clusters identified by the Dirichlet Multinomial Mixtures model.

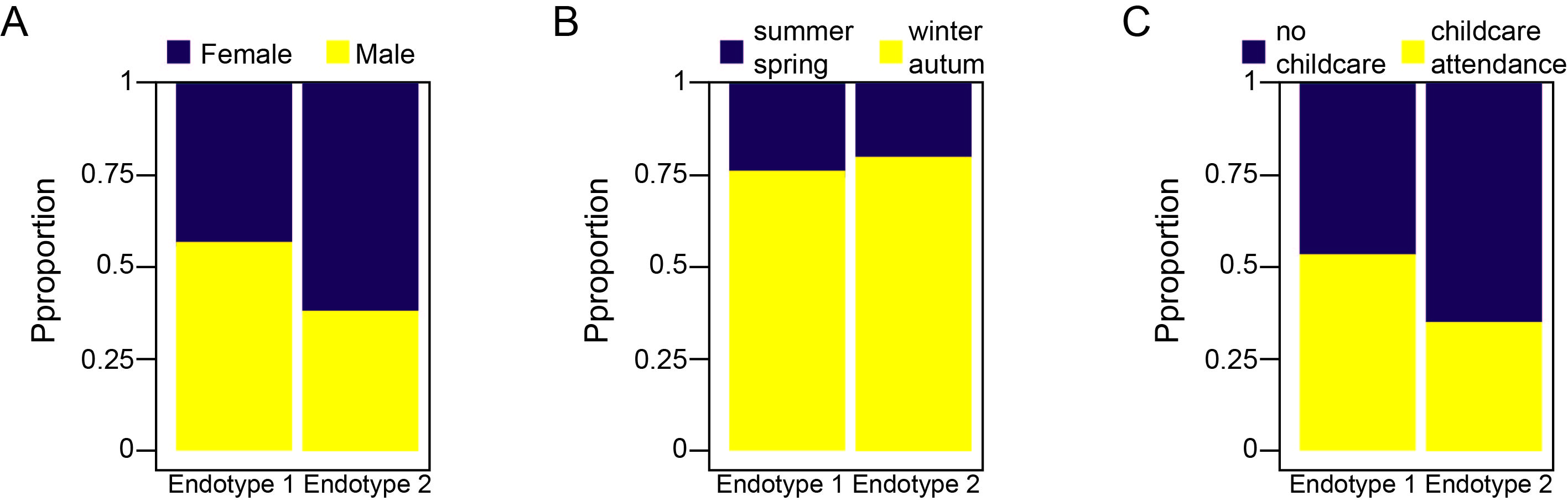

**Figure S9.** Percentage stacked barcharts showing the proportion of samples within each microbial community cluster by the sex of the infants from whom swabs were obtained (**A**), and by the season during which they were collected (**B**). Stacked barcharts in **C** show the proportion of each microbial community type present in background swabs in relation to whether the participant attended childcare. Fisher exact test (FET) indicated no association between endotype (considering both asymptomatic and symptomatic swabs) and sex (**A**, *p*=0.19), or seasonality (**B**, *p*=0.93). Similarly, endotype in asymptomatic swabs was not associated with childcare attendance (**C**, *p*=0.49).

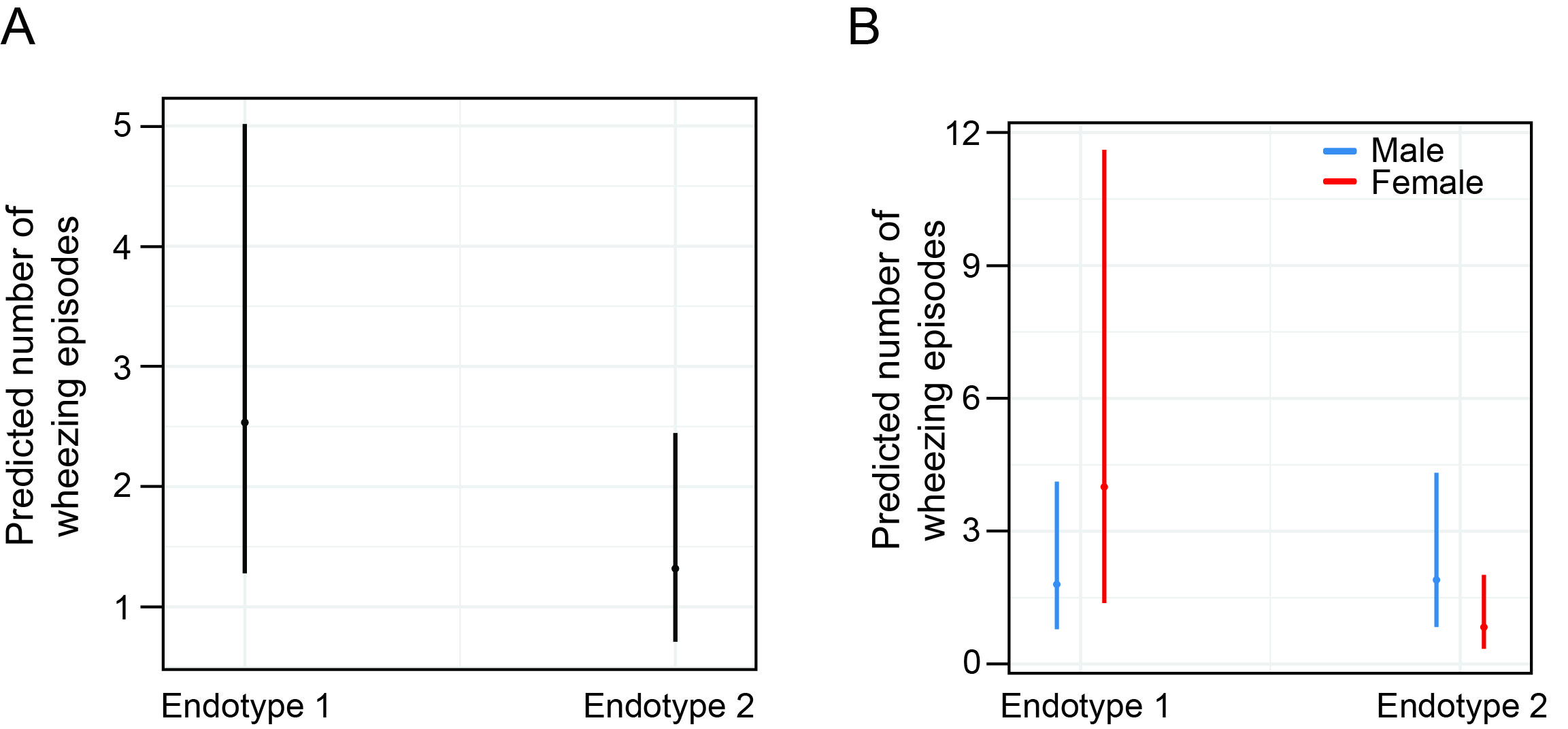

**Figure S10**. Marginal predicted values with 95% confidence intervals from negative binomial regression. Predicted values are shown for: (**A**) number of wheezing episodes, and (**B**) number of wheezing episodes after adjusting the model for sex, during the first year of life, across both microbial endotypes, as estimated using the R function ggpredict.

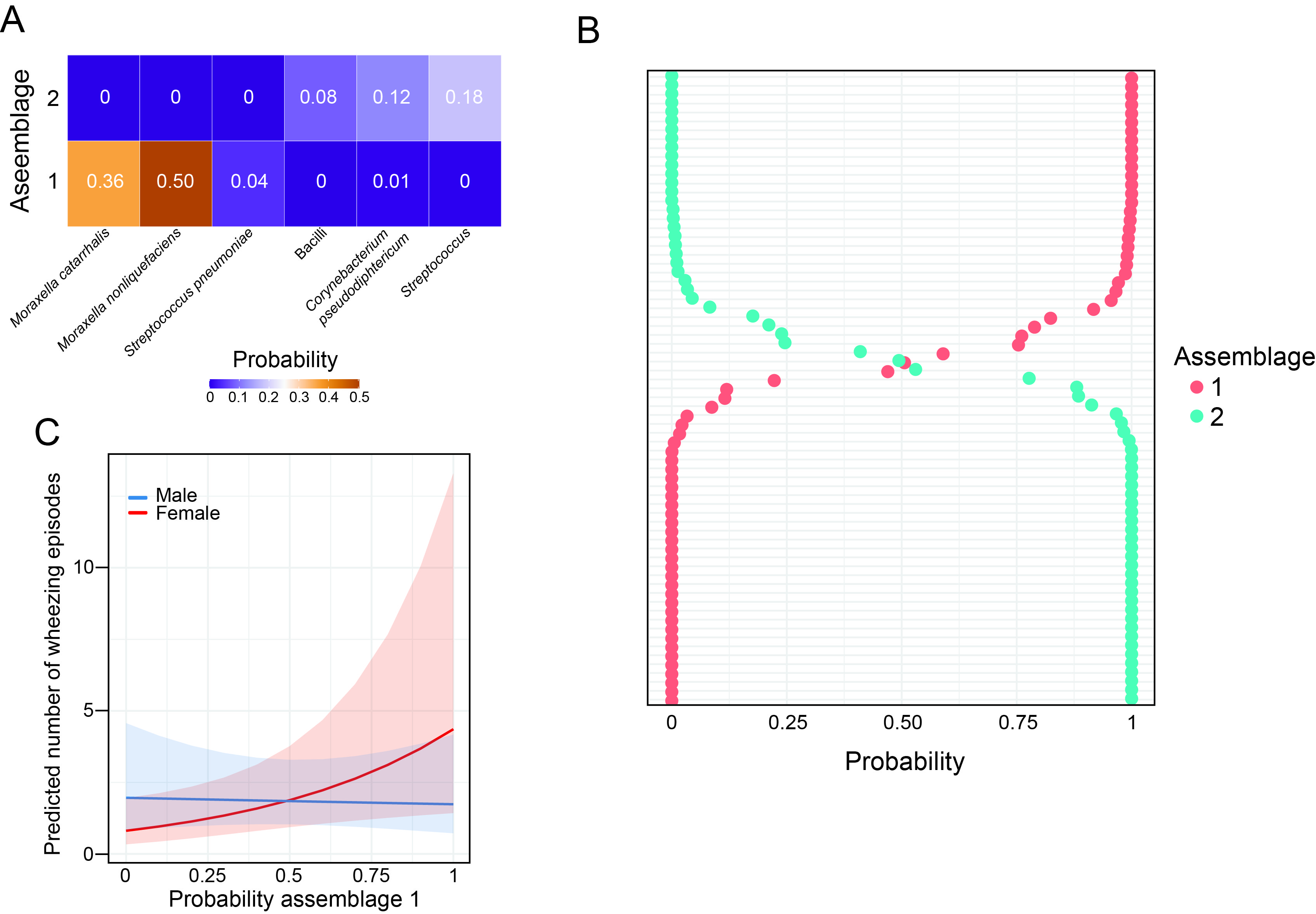

**Figure S11.** Latent Dirichlet Allocation (LDA) was applied to model microbial assemblages based on ASV-level abundance data. LDA is a probabilistic topic modelling approach originally developed for text analysis but increasingly used in microbial ecology to uncover co-occurring taxa (14). CLR transformation was not applied prior to LDA, as the model assumes count-based input. LDA was implemented using the topicmodels package in R (v0.2-17), specifying a Gibbs sampling approach for model fitting. The number of topics (microbial assemblages) was selected based on topic coherence and interpretability. A two-assemblage model was selected based on interpretability and coherence metrics. Each sample (swab-associated bacterial profile) was represented as a probabilistic mixture of topics (assemblages), and each topic (assemblage) was characterized by a probability distribution over taxa. **A.** ASV distribution across the two microbial assemblages. Rows represent assemblages and columns illustrate the three ASVs with the highest probability in each assemblage. Assemblage 1 was characterised by a dominance of ASVs assigned to *Moraxella nonliquefaciens* and *Moraxella catarrhalis*, with relative abundances of 50% and 36% respectively. In contrast, assemblage 2 was primarily defined by moderate abundances of ASVs representing typical bacterial commensals, including *Streptococcus* (18%), and *Corynebacterium pseudodiphteriticum* (12%). **B.** Estimated assemblage probabilities for each swab bacterial profile. Each row represents an individual swab sample and the LDA‑derived probabilities of belonging to assemblage 1 or assemblage 2. **C.** Marginal predicted incidence rates of wheezing episodes by probability of belonging to assemblage 1 adjusted by sex, derived from the negative binomial regression model. Coloured lines show the predicted rates for females (red) and males (blue) across the range of assemblage 1 probability values, and shaded ribbons denote 95% confidence intervals. The model shows a significant sex–assemblage interaction in predicting wheezing frequency. In females, each unit increase in assemblage 1 probability was associated with a higher wheezing rate (*β* = 1.68 ± 0.74, *z* = 2.28, *p* = 0.023), corresponding to a IRR of approximately 5.38. The assemblage 1-by-sex interaction was negative but not significant (*β* = –1.81 ± 0.99, *z* = –1.83, *p* = 0.067), suggesting that the association between assemblage 1 and wheezing may be weaker in males.

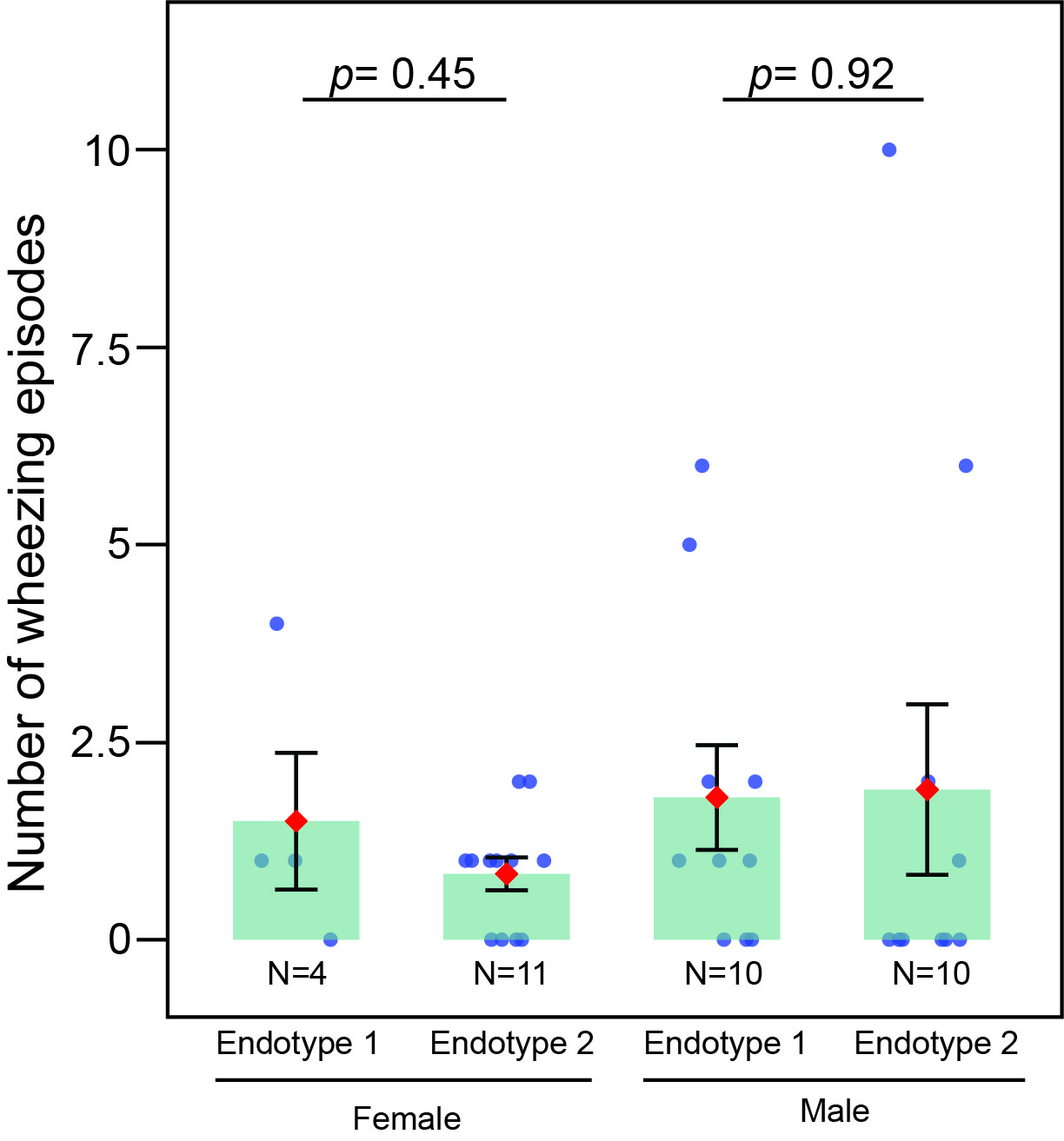

**Figure S12. Figure 3E (sensitivity analysis):** Excluding the high‑leverage sample, observed versus predicted wheezing counts are shown. Grey points represent individual observations, while the bars display the mean observed values per endotype-by-sex group, with error bars indicating the standard error of the mean. Red diamonds indicate model-predicted wheezing episodes. Group sample sizes (N) are shown beneath each bar. Estimated marginal means were compared using Wald z-tests with Dunnett adjustement relative to the reference endotype (endotype 1).

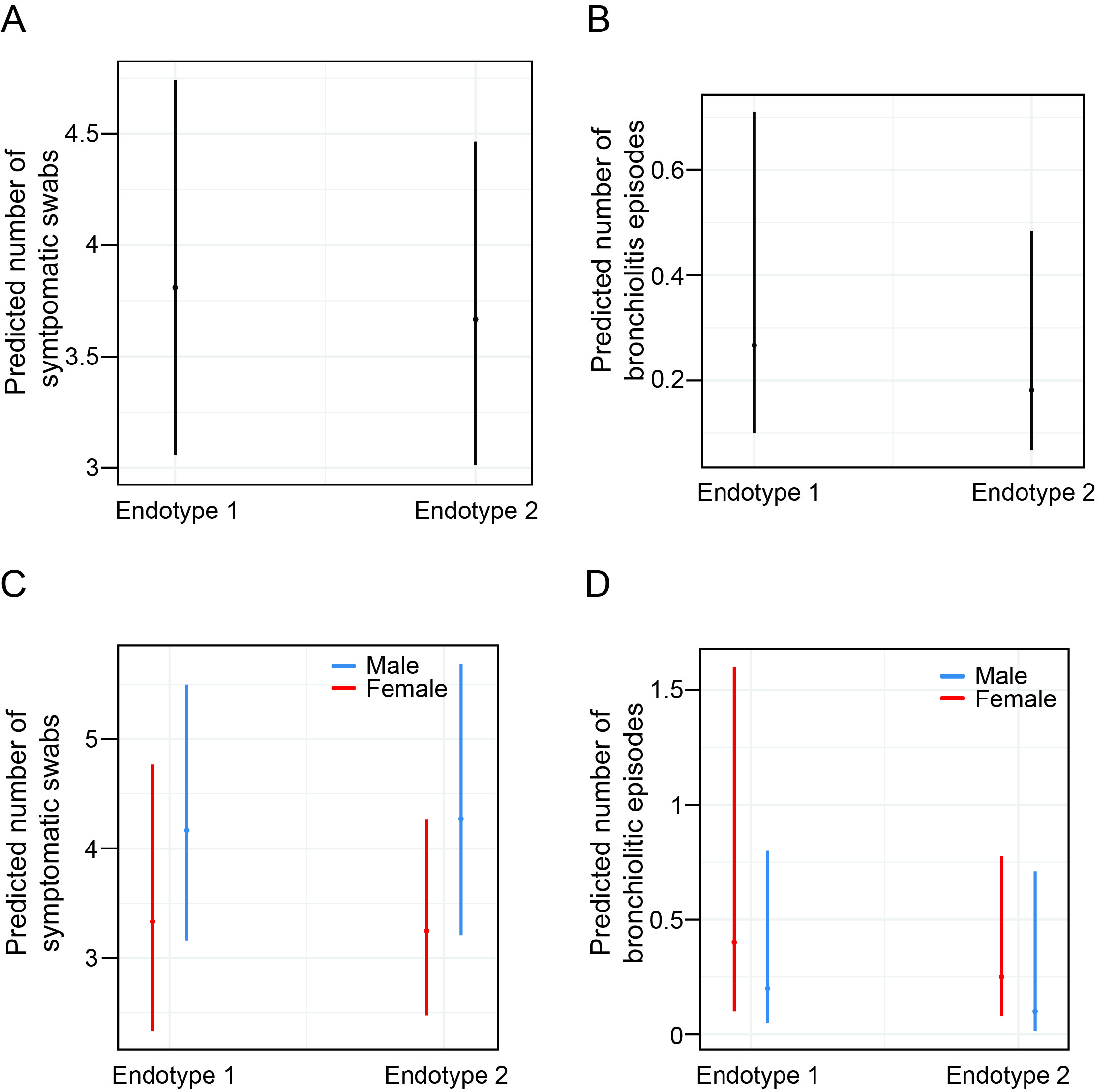

**Figure S13**. Marginal predicted values with 95% confidence intervals from Poisson regression models. Predicted values are shown for: (**A**) number of symptomatic swabs, and (**B**) number of bronchiolitis episodes during the first year of life, across both microbial endotypes, as estimated using the R function ggpredict. Panels (**C**) and (**D**) show predicted values for (**C**) number of symptomatic swabs and (**D**) number of bronchiolitis episodes during the first year of life, with models adjusted for sex.

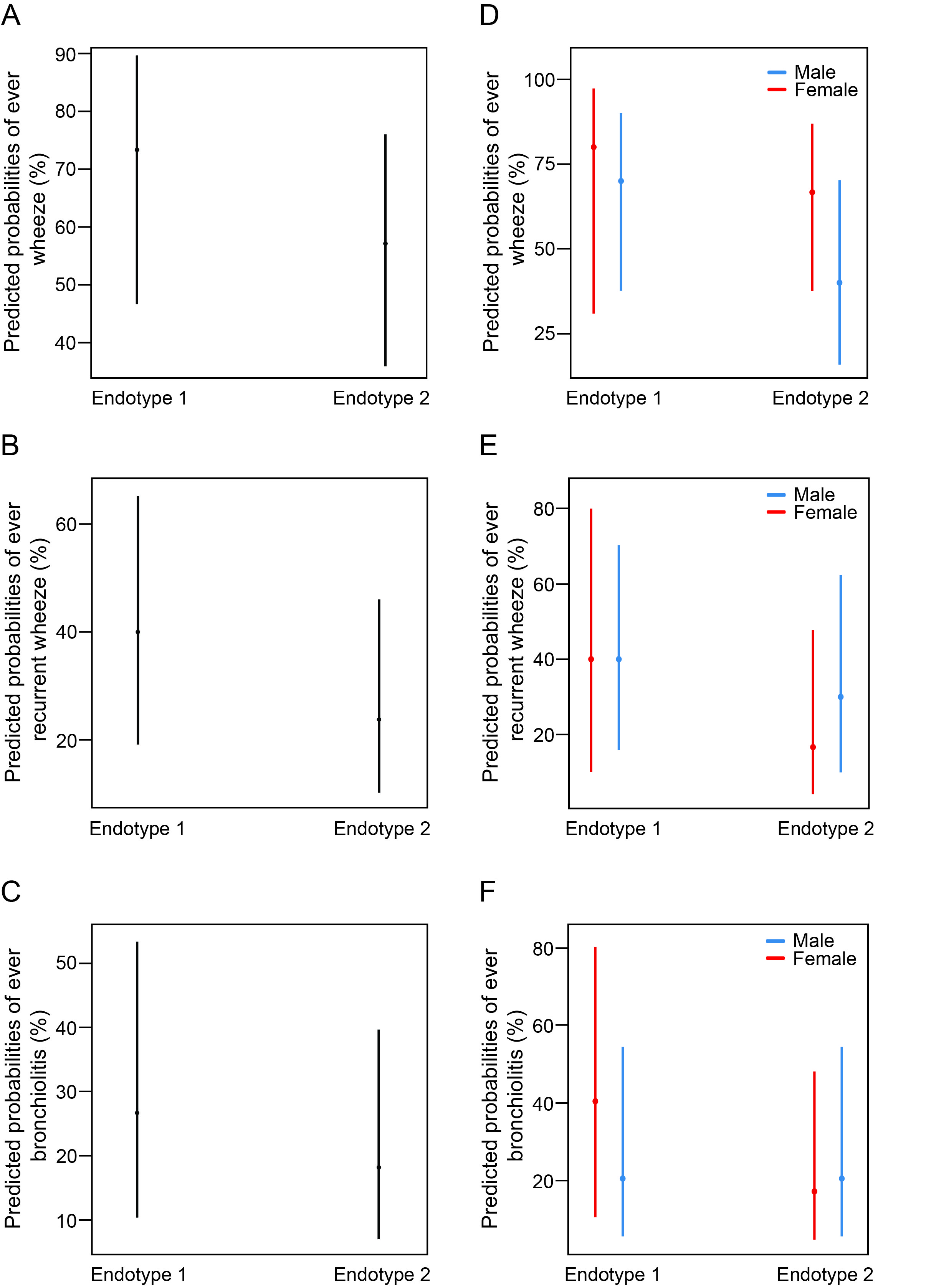

**Figure S14.** Marginal predicted probabilities with 95% confidence intervals from logistic regression models. Predicted probabilities are shown for: (**A**) ever wheeze, (**B**) ever recurrent wheeze, and (**C**) ever bronchiolitis, in the first year of life, across both microbial endotypes, as estimated using the R function ggpredict. Panels (**D-F**) show predicted probabilities with models adjusted for sex.

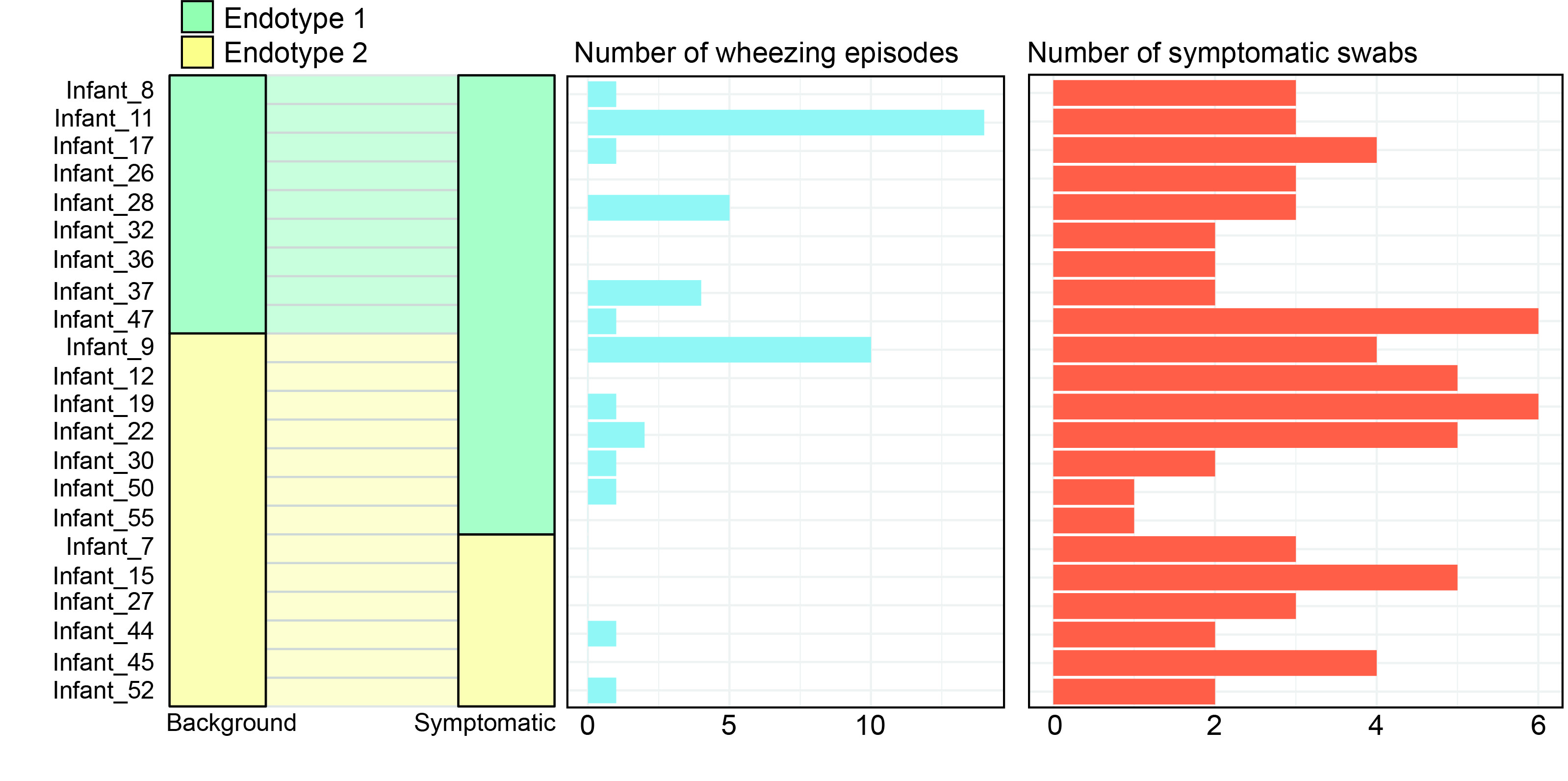

**Figure S15.** The alluvial plot (left) illustrates transitions in microbial endotypes from background to symptomatic collection timepoints. Each participant is represented as a flow from the endotype detected in their background swab to the endotype observed in the subsequent symptomatic swab. The bar plots in the middle and on the right display the number of wheezing episodes and symptomatic swabs, respectively, during the first year of life, with participant ID order consistent across all panels. Three transition patterns were identified during symptomatic swabs: pattern 1 (endotype 1 to endotype 1), pattern 2 (endotype 2 to endotype 1), and pattern 3 (endotype 2 to endotype 2). We did not observe significant differences in the number of wheezing episodes or symptomatic swabs during the first year of life across transition patterns (wheezing episodes: Wilcoxon rank-sum test [WRST], pattern 1 vs. pattern 2, *p* = 0.62; pattern 1 vs. pattern 3, *p* = 0.66; number of symptomatic swabs: WRST, pattern 1 vs. pattern 2, *p* = 0.28; pattern 1 vs. pattern 3, *p* = 0.29).
